# Autoimmune profiling suggests paraneoplastic etiology in pediatric ROHHAD

**DOI:** 10.1101/2021.06.04.21257478

**Authors:** Caleigh Mandel-Brehm, Leslie A. Benson, Baouyen Tran, Andrew F. Kung, Sabrina A. Mann, Sara E. Vazquez, Hanna Retallack, Hannah A. Sample, Kelsey C. Zorn, Lillian M. Khan, Lauren M. Kerr, Patrick L. McAlpine, Lichao Zhang, Frank McCarthy, Joshua E. Elias, Umakanth Katwa, Christina M. Astley, Stuart Tomko, Josep Dalmau, William W. Seeley, Samuel J. Pleasure, Michael R. Wilson, Mark P. Gorman, Joseph L. DeRisi

**Affiliations:** Department of Biochemistry and Biophysics, University of California, San Francisco, CA; Department of Neurology, Harvard Medical School, Boston, MA; Weill Institute for Neurosciences, Department of Neurology, University of California, San Francisco, CA; Department of Neurology, Boston Children’s Hospital, Boston, MA; Otolaryngology Head and Neck Surgery Research Division, Stanford University, Stanford, CA; Chan Zuckerberg Biohub, Stanford, CA; Department of Pulmonary Medicine, Sleep Center, Boston Children’s Hospital, Boston, MA; Division of Endocrinology & Computational Epidemiology, Boston Children’s Hospital, Boston, MA; Department of Neurology, Washington University, St. Louis, MO; Catalan Institution for Research and Advanced Studies (ICREA), Hospital Clinic-Idibaps, University of Barcelona, Barcelona, Spain; Memory and Aging Center, Department of Neurology, University of California, San Francisco, CA; Chan Zuckerberg Biohub, Department of Biochemistry and Biophysics, University of California, San Francisco, CA

**Author notes:** Authors contributed equally to this work (Caleigh Mandel-Brehm, Leslie A. Benson, Baouyen Tran). Co-Corresponding authors **Corresponding Author Information** Name: Mark P. Gorman, Address: 300 Longwood Ave, Boston, MA, 02115, Name: Joseph L. DeRisi, Address: 1700 4^th^ street, QB3 room 404, San Francisco, CA 94110.

## Abstract

ROHHAD (<u>R</u>apid-onset <u>O</u>besity with <u>H</u>ypothalamic Dysfunction, <u>H</u>ypoventilation and <u>A</u>utonomic <u>D</u>ysregulation) is a rare, yet severe pediatric disorder resulting in hypothalamic dysfunction and frequent sudden death. Genetic and other investigations have failed to identify an etiology or diagnostic test. Frequent co-occurrence of neuroblastic tumors (NTs) and cerebrospinal fluid inflammation point to an autoimmune paraneoplastic neurological syndrome (PNS); however, specific anti-neural autoantibodies, a hallmark of PNS, have not been identified. Here, we screened antibodies from a curated cohort of ROHHAD patients (n=9) and controls (n=150) using a programmable phage display of the human peptidome (PhIP-Seq). Our ROHHAD cohort exhibited frequent association with NTs (8/9) and features consistent with autoimmune etiology. Autoantibodies to Zinc finger and SCAN domain-containing protein 1 (ZSCAN1) were discovered and orthogonally validated in 7 of 9 ROHHAD patients, all of whom had NTs, and shown to be absent in non-ROHHAD pediatric patients with NTs. Notably, human ZSCAN1 expression was confirmed in ROHHAD tumor and healthy human hypothalamus. Our results support the notion that tumor-associated ROHHAD is a pediatric PNS, potentially initiated by an immune response to peripheral NT. ZSCAN1 autoantibodies may aid in an accurate diagnosis of ROHHAD, thus providing a means toward early detection and treatment. Lastly, given the absence of the *ZSCAN1* gene in rodents, our study highlights the value of human-based approaches in addition to the classical rodent-based approaches for detecting novel PNS subtypes.

## Main

Rapid-onset Obesity with Hypothalamic Dysfunction, Hypoventilation and Autonomic Dysregulation (ROHHAD) is a poorly understood pediatric syndrome with high morbidity and mortality, distinguished by its unique progression of multi-system derangements^1,2^. Variable degrees of hypothalamic and presumed brainstem dysfunction manifest after age two as obesity, autonomic dysregulation and alveolar hypoventilation with risk of sudden death from respiratory or autonomic dysfunction^1–6^. Neuropsychiatric changes and seizures have been described^7–8^. Detailed longitudinal follow up describing ROHHAD’s natural history is limited^9^. Although incurable, early supportive care including respiratory support for hypoventilation may reduce morbidity^10,11^. Diagnosis remains difficult given lack of biomarkers and overlap with other pediatric disorders including congenital central hypoventilation syndrome (CCHS), Prader-Willi syndrome and other forms of genetic and non-genetic obesity^12–15^. Although recognized as a distinct disorder since 2000, ROHHAD is rare with fewer than 200 reported cases^1,5^.

### The etiology is unclear

Thus far, most etiologic investigations into ROHHAD have focused on genetic susceptibility, without consistent results^16–19^. The presence of neuroblastic tumor (NT) defines a large subset of patients with ROHHAD (30-100% depending on the series), sometimes referred to as ROHHAD-NET^5,20^. NTs are similarly associated with CCHS, a genetic condition, and opsoclonus-myoclonus syndrome (OMS), an autoimmune paraneoplastic neurological syndrome (PNS)^21–23^. With respect to ROHHAD, several observations suggest an immune-mediated disease, including the presence of oligoclonal bands in cerebrospinal fluid (CSF) and immune-cell infiltrates in brain, including hypothalamus and brainstem at autopsy^24–29^. In addition, some patients with ROHHAD have variable improvement with immunotherapy^7,30,31^. Establishing ROHHAD as a PNS would have important implications for developing diagnostic biomarkers, highlighting the need for tumor identification and resection, and advancing the use of immunosuppressive treatments^32^.

On a molecular level, a PNS can be defined by its association with autoantibodies to specific onconeural antigens, or proteins expressed in both the pathogenic tumor and healthy nervous system^33^. Despite a suspicion of PNS in ROHHAD^29,34^, evidence of anti-neural reactivity is limited and an associated onconeural antigen has not been identified. Here, we deployed human-specific phage-display immunoprecipitation and next-generation sequencing (PhIP-Seq) and complementary techniques to investigate a possible paraneoplastic etiology in ROHHAD. We find that autoantibodies to protein Zinc finger and SCAN domain-containing protein 1 (ZSCAN1) are robustly associated with ROHHAD and may be useful as a diagnostic biomarker and advancing knowledge of this poorly understood disease.

## Results

### Clinical profile of ROHHAD cohort

The ROHHAD cohort consisted of 9 patients (ROHHAD-1 through 9) selected on the basis of clinical suspicion for ROHHAD syndrome, without alternative diagnosis (Table1). Each patient was found to exhibit defining features of ROHHAD syndrome including rapid-onset of obesity between 2-5 years of age and multiple additional features of hypothalamic dysfunction (Table1). Three patients were described previously^1,7,35^. Hypoventilation was present in 8/9 at diagnosis. Eight of 9 patients were found to have NT (ROHHAD-1 through 8), while a single patient in this cohort was not known to have a tumor (ROHHAD-9), although work-up was limited to chest x-ray and chest MRI without abdominal imaging. Eight of 9 subjects had clinical genetic testing without genetic syndromes identified to explain their phenotypes. Features suggestive of a neuroinflammatory process include comorbid autoimmune diseases (2/9), elevated CSF neopterin (3/9), marked inflammation noted within resected tumor tissue (4/8) and matched or CSF-specific oligoclonal bands (5/8 tested). Improvement in symptoms was observed in 6/7 patients treated acutely with supportive care, tumor resection and immune therapy at diagnosis. Response to immunotherapy was most clearly demonstrated in ROHHAD-6 in whom CO_2_ retention improved and worsened in parallel with changing steroid doses (Supplemental Figure 1). Similarly, neurobehavioral symptoms improved for ROHHAD-7^6^. Additional clinical details pertaining to autoimmune features and neurologic phenotypes for the ROHHAD cohort are provided in Supplemental Table 1 and Supplemental Table 2, respectively.

**Table 1.** Clinical profile of ROHHAD cohort.

| Patient No. and sex | Sample Type | ROHHAD Symptoms |  |  |  |  | Treatments# | Outcome |
| --- | --- | --- | --- | --- | --- | --- | --- | --- |
|  |  | Rapid-Onset Obesity? | Respiratory phenotype | Endocrinopathies† | Autonomic Dysfunction | Tumor Type and CSF Results |  |  |
| 1 (F) | CSF, Serum | Yes | CSA, 24 hour hypoventilation | Thirst dysregulation, AVP dysregulation (with DI and SIAD), hyperprolactinemia, HH, GHD, CH, CAI, obesity (Z=+2.5, peak +2.7), IR, T2D (metformin, insulin, GLP1RA) | Temperature and heart rate dysregulation, enuresis, duodenal pseudoobstruction | Ganglioneuroma; 1 white blood cell/m<sup>3</sup>, 8 red blood cells/m<sup>3</sup>, 0 OCBs | Tumor resection, trach/vent, IVIG, rituximab, MMF | Alive |
| 2 (F)<sup>35</sup> | CSF, Serum | Yes | Night-time hypoventilation | Hyperprolactinemia, HH, GHD, obesity (Z=+2.4, peak +3.1), IR, prediabetes | Constipation, encopresis | Ganglioneuroma; 0 white blood cell/m<sup>3</sup>, 54 red blood cells/m<sup>3</sup>, matched CSF and serum OCBs | Tumor resection, CPAP, IVIG, Rituximab | Alive |
| 3 (M) | CSF, Serum | Yes | CSA, 24 hour hypoventilation | Thirst dysregulation, AVP dysregulation (with DI and SIAD), prepubertal gonadotropins, GHD, CH, CAI (partial), obesity (Z=+2.6, peak +5.2) | Temperature instability, heart rate variability, severe constipation with rectal prolapse, episodic diaphoresis | Neuroblastoma; 5 white blood cell/m<sup>3</sup>, 525 red blood cells/m<sup>3</sup>, 13 OCBs at diagnosis | Tumor resection, trach/vent, IVIG, rituximab | Sudden death |
| 4 (M) | CSF, Serum, Tumor | Yes | CSA, 24 hour hypoventilation | Hyperprolactinemia (resolved), obesity(Z=+3.1, peak +3.7) | Hypertension, severe paroxysmal sympathetic hyperactivity at times with flash pulmonary edema, (GI dysmotility post arrest,<br>improved) | Ganglioneuroblastoma; 4 white blood cell/m<sup>3</sup>, 2 red blood cells/m<sup>3</sup>, 2 OCBs (normal), neopterin 20 (normal) | Tumor resection, trach/vent, methylprednisolone, IVIG, rituximab | Alive |
| 5 (M) | CSF,<br>Serum,<br>Tumor | Yes | CSA, 24 hour<br>hypoventilation | SIAD, hyperprolactinemia,<br>CH, obesity (Z=+4.7, peak<br>6.1) | Blood pressure,<br>heart rate and<br>temperature<br>instability (severe<br>hyperthermia up to<br>108F w/ brain<br>injury), episodic<br>diaphoresis | Ganglioneuroblas<br>toma; 0 white<br>blood cell/m3, 0<br>red blood<br>cells/m3, 0<br>OCBs, neopterin<br>41 (normal)<br>(repeat LP after<br>5.5 months<br>similar now with<br>matched CSF<br>and serum<br>OCBs) | Tumor<br>resection,<br>trach/vent,<br>dexamethason<br>e, IVIG,<br>rituximab, high<br>dose<br>cyclophospha<br>mide | Alive,<br>severe<br>debilitatio<br>n and<br>behavior<br>problems |
| 6 (M)** | CSF,<br>Serum | Yes | 24 hour<br>hypoventilation | Thirst dysregulation, AVP<br>dysregulation (with DI),<br>hyperprolactinemia, CH,<br>obesity (Z=+1.7, peak +3.9) | Transient<br>temperature<br>instability at<br>diagnosis,<br>episodic<br>diaphoresis (all<br>resolved with<br>treatment) | Ganglioneuroblas<br>toma ,<br>intermixed; 36<br>white blood<br>cell/m3, 0 red<br>blood cells/m3, 0<br>OCBs, neopterin<br>94 (elevated) | Tumor<br>resection,<br>BiPAP,<br>dexamethason<br>e, IVIG,<br>rituximab | Alive |
| 7 (F)<sup>7</sup> | Serum | Yes | OSA | DI (partial) | None | Ganglioneuroblas<br>toma | Rituximab,<br>high dose<br>cyclophospha<br>mide | Alive |
| 8 (F)<sup>1</sup> | CSF,<br>Serum | Yes | 24 hour<br>hypoventilation | Mild hyperprolactinemia,<br>Hypothyroidism, central<br>precocious puberty, later<br>amenorrhea, GHD,<br>obesity (BMI 38.7 kg/m2<br>adulthood), recurrent<br>hypoglycemia/hypokalemia<br>in adulthood of unclear<br>etiology | None<br>diagnosed/evaluat<br>ed in childhood | Ganglioneuroma<br>; 2 white blood<br>cell/m3, 60 red<br>blood cells/m3,<br>protein 141, 0<br>OCBs, neopterin<br>21 (normal) | Steroids, IVIG,<br>rituximab,<br>thymectomy | Alive |
| 9 (M) | Serum | Yes | CSA, 24 hour hypoventilation | Hypodipsia, AVP dysregulation (with DI and hyponatremia), obesity (Z=+2.7, peak +4.7), IR, T2D (metformin, insulin) | Temperature instability, bradycardia, hypothermia, episodic diaphoresis | None (abdomen not imaged); 2 white blood cell/m3, 31 red blood cells/m3, 1 matched serum to CSF OCB, neopterin 76 (elevated) | IVIG | Sudden death |

AVP = Arginine Vasopressin, BiPAP= Bilevel Positive Airway Pressure, BMI = Body Mass Index, CAI=central adrenal insufficiency, CH=central hypothyroidism, CSF = Cerebrospinal Fluid, CPAP= Continuous Positive Airway Pressure, CSA= central sleep apnea, DI= diabetes insipidus, GHD=growth hormone deficiency, GLP1RA = Glucagon-like peptide-1 receptor antagonists, LP= Lumbar Puncture, HH=hypogonadotropic hypogonadism, IR=insulin resistance, IVIG = Intravenous Immunoglobulin, OCB = Oligoclonal Band, MMF = Mycophenolate Mofetil, OSA = Obstructive Sleep Apnea, SIAD=syndrome of inappropriate antidiuresis, T2D=type 2 diabetes mellitus, trach=tracheostomy, vent = ventilator
\* All tested CSF had normal protein, glucose and IgG index values except where noted
\*\*See Supplemental Figure 1

### Clinical profile of control cohorts

Control cohort 1 (CC1) and control cohort 2 (CC2) were assembled for PhIP-seq screening and validation experiments, respectively. CC1 represents plasma from 100 de-identified blood donors courtesy of New York Blood Center. CC2 represents 27 de-identified healthy volunteers (plasma n =3; sera n = 17, CSF n =7) and 26 clinically relevant pediatric controls, Pediatric controls include: childhood obesity + NT (n =1), and children with OMS (with NT n = 10, without NT n=15). Clinical details and demographics pertaining to controls is provided in Supplemental Table 3 (CC1) and Supplemental Table 4 (Pediatric controls OMS and Obesity +NT).

### Screening for autoantibodies using rodent based approaches

Immunohistochemical screening of patient antibodies against neural tissue, typically rodent, is a classical approach for identifying cryptic autoimmune etiologies among idiopathic neurological syndromes^36–40^. In two independent laboratories (JLD, SJP), antibodies in CSF from ROHHAD-1 through 6 were tested for immunoreactivity against mouse brain tissue (Supplemental Methods). Neither identified anti-neural reactivity in any patient (Supplemental Figure 2). Additional, independent assessment of autoantibodies to extracellular neural targets were also tested in 4 patients in this cohort using pre-established protocols for fixed^39^ and live rat neurons^40^. All studies were negative (JD, see supplemental Table 1).

### Screening for autoantibodies using human-specific Phage-Display

#### Immunoprecipitation and Sequencing (PhIP-Seq)

The PhIP-seq library employed here displays a representation of the human proteome and has been used previously for detecting diagnostic PNS autoantibodies ^40–41^. As part of a discovery pilot, we screened CSF from 3 ROHHAD patients (ROHHAD 1-3) and plasma from CC1 (n=100). Individual PhIP-seq datasets were normalized to 100,000 reads (RP100K) and protein RP100Ks were averaged according to cohort. To aid in candidate identification, ROHHAD-specific Z-score enrichments were generated, using CC1 values (see methods). All proteins with a ROHHAD mean RP100K > 0 were plotted relative to Z-score enrichments (Figure 1). Candidate antigens were expected to satisfy the following criteria in at least 2 patients: a minimum level of recovery (RP100K>50) and minimum Z-score enrichment (>3). Of the ∼20,000 protein possibilities, only 1 protein, ZSCAN1, satisfied these criteria. Using the same criteria above, enrichment of ZSCAN1 was not observed in other previously published autoimmune cohorts screened on the same PhIP-Seq platform including PNS syndromes anti-Yo (n= 36)^41^, anti-Hu (n= 44)^41^, anti-Ma2 (n =2)^42^, anti-KLHL11 (n =7)^42^ or patients with systemic Autoimmune Polyglandular Syndrome Type I (n=39)^43^.

**Figure 1:**
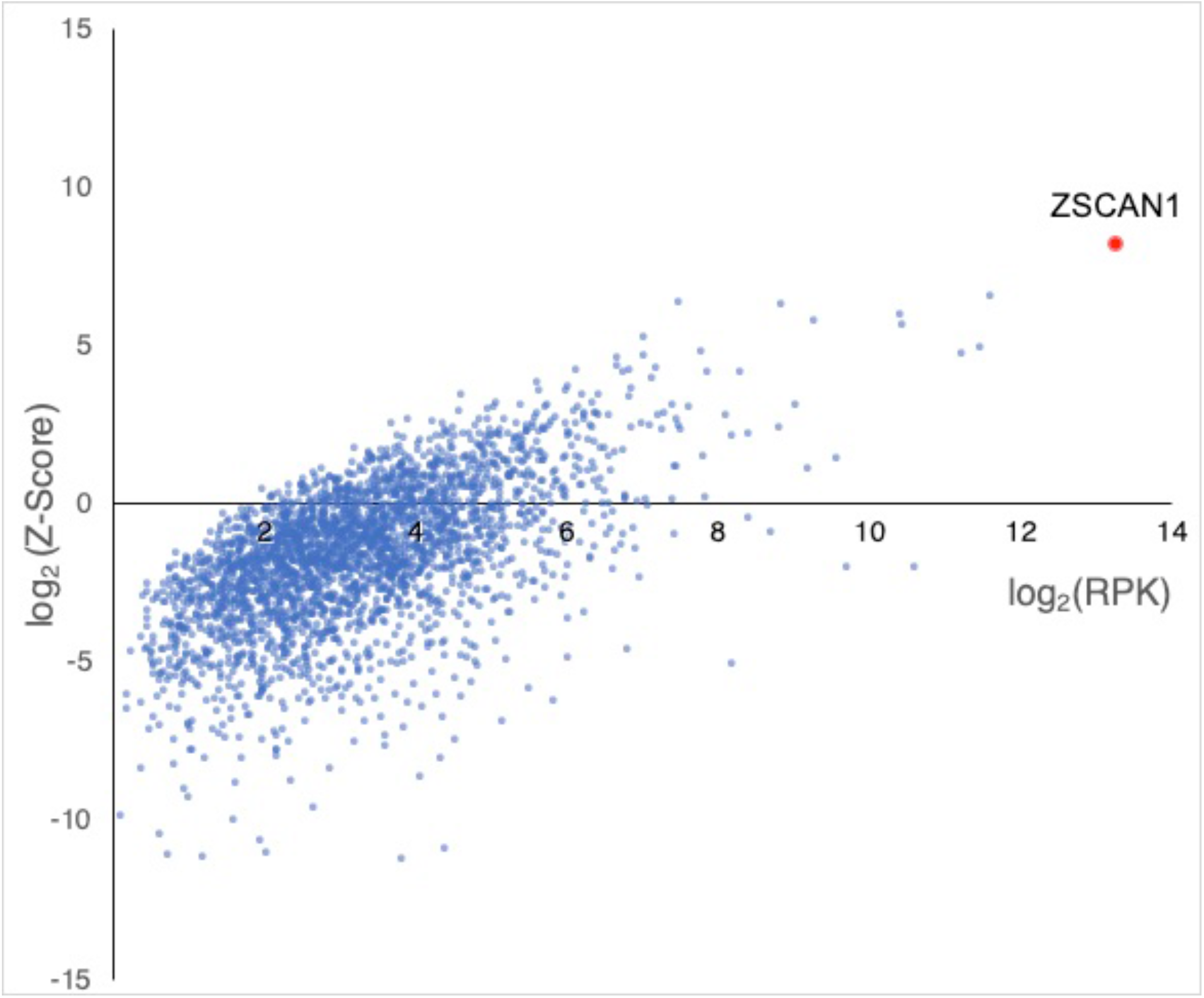
PhIP-Seq screen implicates ZSCAN1 as a candidate autoantigen in ROHHAD. CSF from 3 ROHHAD patients (ROHHAD-1 to 3) and plasma from a large set of “healthy controls” (n = 100) were tested by PhIP-Seq. Individual data was averaged according to cohort. All proteins with a ROHHAD mean RP100K >0 are plotted against ROHHAD Z-score enrichments compared to healthy. Non-ZSCAN1 proteins are denoted with blue dots, ZSCAN1 is denoted with a red dot.

#### Orthogonal validation assays

To further investigate the association of ZSCAN1 and ROHHAD syndrome, we repeated PhIP-seq with an expanded ROHHAD cohort (including the 4 original patients) and new set of controls (CC2). For all samples, ZSCAN1 RP100K is reported, as well as, Z-score enrichment based on our background control dataset CC1 (ZSCAN1 mean = 8.18 RP100K, standard deviation = 32.53 RP100K), with Z-score > 3 considered positive. We reproduced the association between ZSCAN1 and ROHHAD, showing enrichment in 7 of 9 ROHHAD patients (ROHHAD-1 to 7) and 0 of 50 samples from CC2 (Figure 2, *top*). We next leveraged peptide-level data and found antigenicity within ZSCAN1 was largely restricted to the C-terminal of ZSCAN1, with patients displaying enrichment of overlapping peptides, increasing confidence in our findings (Supplemental Figure 3). Intriguingly, patients ROHHAD-1 through 7 shared a common 11 amino acid (AA) epitope.

**Figure 2.**
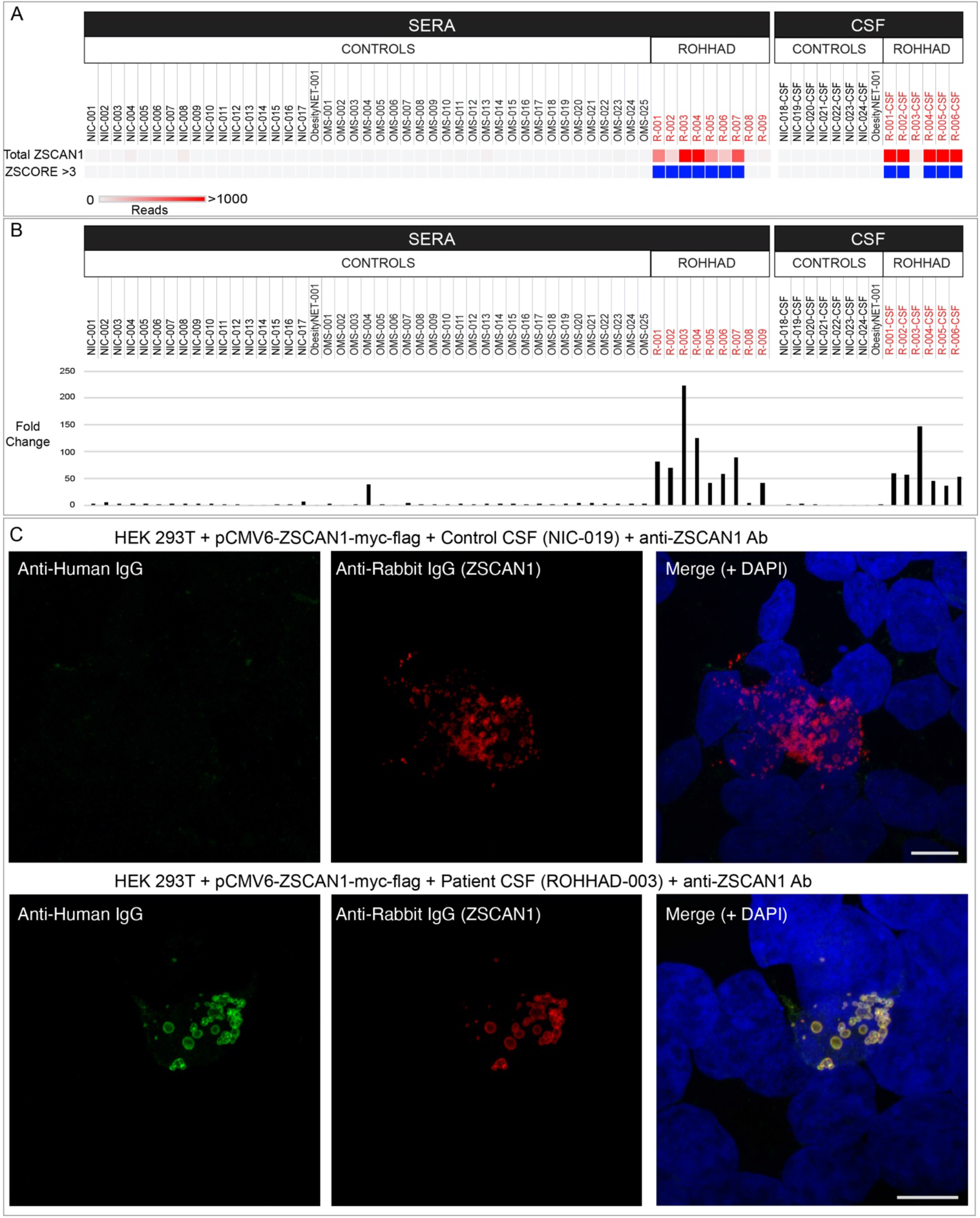
Validation of autoantibodies to ZSCAN1 in ROHHAD patients. In panel **a** and **b**, enrichment of ZSCAN1 by PhIP-Seq and Radio Ligand Binding Assay (RLBA) was compared between ROHHAD patients (n=9), non-inflammatory healthy controls (n=24) and pediatric controls including children with OMS with and without NT (n=25) and an obesity patient with NT (n=1). Data represents the average of two independent technical replicates. **a**, PhIP-Seq analysis. Each column represents an individual sample. A heatmap of total ZSCAN1 RP100K is shown in the top row. To enable comparisons between the majority of samples with lower signal we added a ceiling value of 1000 RP100K. Z-score enrichments based on our 100 human donor dataset are plotted below, with samples that have Z-score enrichments > 3 colored with blue squares. Grey squares indicate Z-score < 3. **b**, RLBA testing immunoprecipitation of recombinant ZSCAN1 by ROHHAD patients and controls. For all samples, fold change was calculated by dividing by the mean value from control sera (n=17, mean = 20.83). **c**, Representative image showing immunostaining of 293T cells expressing full-length ZSCAN1 with human CSF and commercial antibody to ZSCAN1 (Rabbit). Top row shows immunostaining with control CSF. Bottom row shows staining with ROHHAD-3 CSF. Colocalization is indicated by yellow in the merge images (far right both rows).

To confirm ZSCAN1 enrichment in our ROHHAD cohort was antibody-mediated, we tested for IP of recombinant ZSCAN1 in two orthogonal assays, including a radioligand binding assay (RLBA) and 293T cell-based overexpression assay (CBA). For the 6 patients with paired CSF and sera (ROHHAD 1-6), ZSCAN1 enrichment in one sample type was sufficient to be called positive in a given assay. First, the RLBA with *in vitro* transcribed and translated full length ZSCAN1 protein revealed positive IP (fold change > 10) by ROHHAD patients 1 through 7 (sera n = 7/7; paired CSF n = 6/6) and no enrichment in CC2 healthy controls (Figure 2B). Low-level enrichment of ZSCAN1 was observed in 2 patients negative by PhIP-seq; ROHHAD-9 and OMS-4. Second, using immunocytochemistry and 293T cells expressing full-length ZSCAN1, we show colocalization of human antibodies with commercial antibody to ZSCAN1 for ROHHAD patients 1 through 7 (sera n=2 ROHHAD-1 and 3; CSF n = 6 ROHHAD-1 through 6). Co-localization is not observed for ROHHAD-8, ROHHAD-9 or controls, including OMS-4 (Figure 2C). Taken together, ROHHAD-1 through 7 reproduced ZSCAN1 enrichment in two antibody-based orthogonal assays, thus limiting possibility of false-positives and validating autoantibodies to ZSCAN1 in this cohort^44^.

Of note for 293T CBAs, we observed increased sensitivity to ZSCAN1 autoantibody detection using CSF compared to sera. This is exemplified best in data collected from the 6 patients with paired CSF and sera (ROHHAD-1 through 6). In 293T CBAs, ZSCAN1 autoantibodies were detected using 6/6 CSF samples (Supplemental Figure 4). In contrast, only 2/6 paired sera were positive (Supplemental Figure 5). To rule out technical artifacts, sera samples were re-tested on CBA at several dilutions 1:100, 1:1000, 1:2000 and 1:5000, with no change in seronegativity. We further tested for recognition of ZSCAN1 autoantibodies using whole cell lysates prepared from 293T cells overexpressing ZSCAN1. When western blotting denatured lysates with CSF and sera we again observed positive signal with 6/6 CSF but only 2/6 paired sera (Supplemental Figure 6).

#### Testing antigen expression in tumor and brain

Typically, autoantibodies associated with PNS target antigens that are expressed in relevant tumor and healthy brain, otherwise known as onconeural antigens^38^. To test for tumor expression of ZSCAN1, one NT from ROHHAD-3 was sectioned and immunostained with a previously validated commercial antibody to ZSCAN1 (Supplemental Figure 4 through 6). A rabbit secondary only control was used to test for nonspecific staining of Rabbit IgG to infiltrating human IgG and Fc proteins in the tumor. ZSCAN1 expression was apparent in the ROHHAD-associated tumor (Figure 3). To test for hypothalamic expression of ZSCAN1, mass spectrometry analysis was performed using immunoprecipitation from human hypothalamic lysates using commercial antibodies to ZSCAN1. ZSCAN1 peptides were readily detectable, consistent with protein expression in this tissue (Supplemental Table 5).

**Figure 3.**
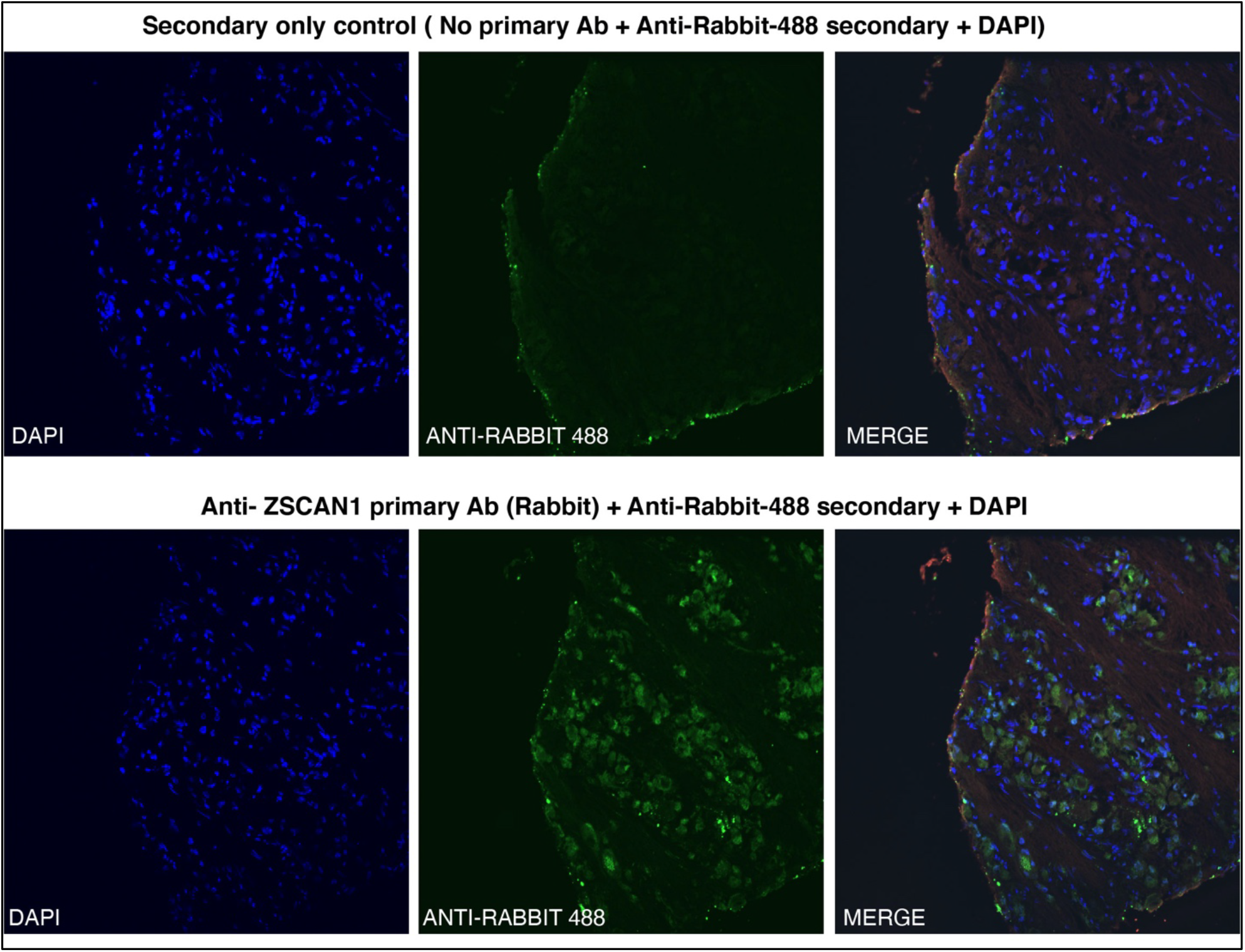
Immunohistochemical detection of ZSCAN1 in NT associated with ROHHAD patient-3. Fixed NT tissue was immunostained with either (top) Anti-Rabbit-488 secondary alone or (bottom) primary antibody to ZSCAN1 (Rabbit) and Anti-Rabbit-488 Secondary. Green: Anti-Rabbit-488 secondary, Blue: DAPI to identify nuclei.

## Discussion

In this work, we describe a proteome-wide screen for autoantibodies in patients with ROHHAD, a complex, diagnostically challenging syndrome with severe and often life-threatening symptoms^1,2^. By comparing antibody profiles from ROHHAD patients to a large set of clinically relevant controls, we found autoantibodies to ZSCAN1 are a putative biomarker of ROHHAD, and thus have potential utility in identifying ROHHAD earlier in the disease course.

All patients in this ROHHAD cohort (n= 9/9) exhibited the defining symptoms of the syndrome including rapid-onset obesity and hypothalamic dysfunction, with an onset between two to five years of age^1,2^. NTs were present in 8/9 patients. Autoantibodies to ZSCAN1 were identified in 7 of 9 patients tested (sera n=7/9, paired CSF n=5/6) by PhIP-Seq, all of whom had NTs. Each of the 7 PhIP-Seq positive samples (ROHHAD-1 through 7) were validated by at least two orthogonal assays, including 293T CBA and RLBA. Two assay validation limits the possibility of false-positive results and is increasingly accepted as a prerequisite for defining bona fide autoantigens^36,44^.

Two ROHHAD patients (ROHHAD-8 and 9) were negative for ZSCAN1 autoantibodies, although CSF was not available for testing. Interestingly, ROHHAD-8 was in clinical remission when their sera was collected and at an older age compared to other patients (age range = 2 – 10 years, median 3 years), who all exhibited active symptoms at sample collection. ROHHAD-9 did not have a tumor identified but exhibited the ROHHAD phenotype and active disease. Future experiments to determine the association of ZSCAN1 antibodies and ROHHAD syndrome in the absence of NT are warranted.

Previous clinical reports suggested ROHHAD could be a PNS by virtue of its co-occurrence with a tumor type already associated with a different PNS (OMS), clinical features of autoimmunity and the failure to identify consistent genetic associations^3,7,8,16–19,24,26–31^. Our findings collectively support the notion of a PNS etiology. First, autoantibodies to the molecular antigen (ZSCAN1) were found in the majority (7/9) of ROHHAD cases, and in all from cases with a NT identified. Second, ZSCAN1 expression in human hypothalamus is consistent with the target tissues affected in ROHHAD patients. Third, ZSCAN1 was found to be expressed in NT tissue from a patient with ROHHAD. Lastly, the presumed intracellular localization of ZSCAN1 (www.uniprot.org, www.proteinatlas.org), together with our expression in ROHHAD NTs as well as healthy nervous system tissue, is reminiscent of other classical PNS onconeural antigens. Antibodies to intracellular onconeural antigens are a biomarker for a major subtype of PNS, such as cerebellar degeneration-related protein 2-like in anti-Yo PNS and neuronal ELAV-like proteins 2,3,4 in anti-Hu PNS^33^. In these cases, autoantibodies themselves are not thought to be directly pathogenic, and the diseases are often less consistently responsive to tumor resection and immunotherapy compared to PNS with antibodies to extracellular antigens. Consistent with the hypothesis of PNS, ROHHAD patients have variable responses (duration and magnitude) to immunotherapy or tumor resection and may respond better to earlier, more aggressive regimens^3,7,30,31^.

The ZSCAN1 protein is a putative zinc finger transcription factor (znTF) that contains a single SCAN domain and 3 Cysteine_2_-Histidine_2_ (C_2_H_2_) zinc finger domains (uniprot.org). Despite nearly equal representation of SCAN domain (84 AA) and C_2_H_2_ domains (100 AA), antigenicity in ZSCAN1 is biased to the C_2_H_2_ domains, a functional region for DNA and RNA binding^45^. Further, although ∼ 700 human proteins containing C_2_H_2_ domains with the motif C-X_2-4-_C-X_12-_H-X_2-6-_H (where X is any AA) are present in our PhIP-seq library, no other ZnTF’s were significantly enriched in more than one patient (Z-score > 1), highlighting the specificity of the motif identified by PhIP-Seq. Importantly, the *ZSCAN1* gene lacks a genetic ortholog in rodents, including mice and rats, with evolutionary divergence towards primate-specificity in the C_2_H_2_ region^45–48^. These observations suggest the putative epitope within ZSCAN1 has a high likelihood of being exclusive to primate. To our knowledge, a primate-specific epitope in autoimmune disease has not yet been described.

A common autoantibody among ROHHAD patients suggests a route to a molecular diagnostic. Here we demonstrate the value of the human-specific PhIP-seq approach for ROHHAD biomarker discovery. *ZSCAN1* has no genetic ortholog in rodents, so the classical rodent based approach for PNS autoantibody detection failed. Our results suggest CBAs or RLBAs on CSF may be sufficient to detect ZSCAN1 autoantibodies in a clinical lab setting. In the case of ROHHAD, these results also suggest that CSF may be the most sensitive sample type for testing, since ZSCAN1 was not always detectable in sera, particularly on CBA, as is true for many PNS.

To summarize, we provide a robust finding of autoantibodies to ZSCAN1 as a marker for tumor-associated pediatric ROHHAD. ZSCAN1 expression in tumor and human hypothalamus provides further evidence to support the clinical suspicion that ROHHAD is a novel type of PNS. This is the first identified intracellular antigen in a PNS unique to children. Further experiments are required to test the utility of ZSCAN1 autoantibodies for diagnosis in ROHHAD patients with and without tumors, define the clinical spectrum of PNS ROHHAD and to understand how immune targeting of ZSCAN1 contributes to the dramatic clinical complications seen in patients with ROHHAD syndrome.

## Methods

All relevant ethical regulations were followed regarding animal experiments and human research participant involvement. We have obtain informed consent from all participants and/or their parents.

### Patient recruitment

All local IRB regulations were followed at Boston Children’s Hospital (protocol #09-02-0043) and University of California, San Francisco (protocol #13-12236). All patient specific data was extracted by chart review (LB, LK, ST) and samples obtained following family consent. ROHHAD subjects were identified by pediatric neuro-immunologists (L.B. or S.T.) on the basis of clinical suspicion for ROHHAD syndrome, without alternative diagnosis.

### Autoantibody discovery and validation

For autoantibody discovery, we utilized the PhIP-Seq library and experimental protocol as previously described.^42^ PhIP-seq datasets were analyzed with a formalized analysis pipeline (generated in-house) that includes alignment of DNA reads to input library^49^, conversion to peptide reads, summing of peptides by protein ID, normalization to 100,000 reads (RP100K) and calculating mean RP100K for each protein. Alignment of sequencing reads to the input library on a peptide-level was performed with RAPsearch2^49^ and command line ‘ /data/bin/rapsearch -d input_library -q read1_location -o output_location -z 12 -v 1 -b 0’. A Z-score enrichment for each protein was calculated based on mean RP100K generated from a large set of healthy controls. Z-score formula = (protein_X_ mean RP100K_Experimental_ – protein_X_ mean RP100K_control_)/ (Standard Deviation of protein_X_ in controls).

### Onconeural antigen investigation

To examine candidate autoantigen expression in ROHHAD tumor and healthy human brain we used immunohistochemistry and immunoprecipitation-mass spectrometry (IP-MS), respectively. For tumor examination, fresh frozen tumor was embedded, sectioned, fixed on slides, stained and imaged according to standard procedures (Supplemental Methods). To detect ZSCAN1 we immunostained with commercial antibody to ZSCAN1 (Rabbit, Thermo Fisher Scientific, PA552488) and anti-rabbit secondary. For IP-MS experiments, ∼100 milligrams of frozen human hypothalamus was homogenized and prepared for IP and subsequent MS analysis (Supplemental Methods). For IP, lysate was diluted to 0.5 mg/mL and one of the following antibodies was added at 1ug/mL: Rabbit anti-ZSCAN1 (Thermo Fisher Scientific, PA552488), Rabbit anti-ZSCAN1 (Sigma Aldrich HPA007938), Rabbit IgG control (Thermo Fisher Scientific, 31235). Positive identification of ZSCAN1 required detection with two commercial antibodies and absence from isotype control. Two technical replicates were performed.

### Animal Protocol

All animal protocols were in accordance with the regulations of the National Institute of Health and approved by the University of California San Francisco Institutional Animal Care and Use Committee (IACUC). Wildtype C57B6 mice were obtained from Jackson Laboratories (Bar Harbor, ME, USA). Adult mice (> postnatal day 42) were used for immunohistological screening of human patient antibodies.

## Data Availability

Data available within the article or its supplementary materials. Raw data files or original microscopy images available upon request.

## Data availability

Original data contributing to the main findings in our study is provided on Dryad repository (temporary link for review: https://datadryad.org/stash/share/-vpGClCcsEFX1bVTwmPAqh3K_6errf8vkOo6k3vPnUk). The dryad repository includes PhIP-Seq analysis and RLBA analysis of ZSCAN1 (Figure 1, Figure 2 a,b), Mass Spectrometry search results (Supplemental Table 5), original microscopy images (Figure 2c, Supplemental Figure 2,4,5), original blots (Supplemental Figure 6). There are no data restrictions for this study.

## Additional Methods

Full experimental details regarding PhIP-Seq screening for candidate autoantibodies, orthogonal validation assays, indirect immunofluorescence screening for anti-neural antibodies on rodent tissue, immunohistochemistry of ROHHAD tumor tissue and IP-MS of human brain are provided in Supplemental Methods.

## Acknowledgements

This research was supported by ROHHAD Fight, Inc, the OMS Life Foundation, the Repository Core for Neurological Disorders, Department of Neurology, Boston Children’s Hospital, and the IDDRC (NIH P30HD018655) and NIH R01MH122471. We would also like to acknowledge the New York Blood Center for contribution of healthy control plasma. Instituto Carlos III (ISCIII, PI20/00197 [JD]), and “La Caixa” Foundation (JD). CMA is supported by NIDDK (K23DK120899). Acknowledge the UCSF Nikon Core Facility for microscope imaging equipment and guidance. JDL is funded by a grant from Chan Zuckerberg Biohub. JDL, MRW, SJP and CMB are funded by the National Institute of Mental Health (NIMH) of the NIH (award 1R01MH122471-01). SJP, BT are also funded by the Brain Research Foundation (Scientific Innovations Award). CMB is also funded by The Emiko Terasaki Foundation (Project 7027742 / Fund B73335) and by the National Institute of Neurological Disorders and Stroke (NINDS) of the National Institutes of Health (award 1K99NS117800-01). SEV is funded by the National Institute of Diabetes and Digestive and Kidney Diseases of the NIH (award 1F30DK123915-01).

## Author contributions

CMB, LB and BT contributed equally to this work.

## Notes

### Competing Interest Statement

The authors have declared no competing interest.

### Author Declarations

All local IRB regulations were followed at Boston Children's Hospital protocol #09-02-0043 and University of California San Francisco protocol #13-12236

## References

1. Katz, E.S., Mcgrath, S., Marcus, C.L. Case Reports Late-Onset Central Hypoventilation With Hypothalamic Dysfunction: A Distinct Clinical Syndrome. Pediatr Pulmonol. 68, 62–8 (2000).

2. Ize-Ludlow, D., et al. Rapid-onset obesity with hypothalamic dysfunction, hypoventilation, and autonomic dysregulation presenting in childhood. Pediatrics 120, e179–88 (2007).

3. Ibáñez-Micó, S., et al. Rapid-onset obesity with hypothalamic dysregulation, hypoventilation, and autonomic dysregulation (ROHHAD syndrome): A case report and literature review. Neurologia. 32, 616–622 (2017).

4. Patwari, P.P. & Wolfe, L. Rapid-onset obesity with hypothalamic dysfunction, hypoventilation, and autonomic dysregulation: review and update. Curr Opin Pediatr. 26, 487–92 (2014).

5. Lee, J.M., et al. Rapid-Onset Obesity with Hypoventilation, Hypothalamic, Autonomic Dysregulation, and Neuroendocrine Tumors (ROHHADNET) Syndrome: A Systematic Review. Biomed Res Int 2018, 1–17 (2018).

6. Chew, H.B., Ngu, L.H., Keng, W.T. Rapid-onset obesity with hypothalamic dysfunction, hypoventilation and autonomic dysregulation (ROHHAD): a case with additional features and review of the literature. BMJ Case Rep. 2011 (2011).

7. Jacobson, L.A., et. al. Improved Behavior and Neuropsychological Function in Children With ROHHAD After High-Dose Cyclophosphamide. Pediatrics 138, e20151080–e20151080 (2016).

8. Sethi, K., et al. ROHHADNET Syndrome Presenting as Major Behavioral Changes in a 5-Year-Old Obese Girl. Pediatrics 134, e586–9 (2014).

9. Eldin, A.W.J., et al. Natural history of ROHHAD syndrome: development of severe insulin resistance and fatty liver disease over time. Clin. Diabetes Endocrinol. 5, 9 (2019).

10. Stowe, R.C. & Afolabi-Brown, O.Pulmonary hypertension and chronic hypoventilation in ROHHAD syndrome treated with average-volume assured pressure support. Pediatr. Investig. 3, 253–256 (2019).

11. Selvadurai, S., et al. Sleep-disordered breathing, respiratory patterns during wakefulness and functional capacity in pediatric patients with rapid-onset obesity with hypothalamic dysfunction, hypoventilation and autonomic dysregulation syndrome. Pediatr. Pulmonol. 56, 479–485 (2021).

12. Kocaay P., Şıklar Z., Çamtosun E., Kendirli T., Berberoğlu, M. ROHHAD Syndrome: Reasons for Diagnostic Difficulties in Obesity. J Clin Res Pediatr Endocrinol. 6, 254–257 (2016).

13. Barclay, S.F., et al. ROHHAD and Prader-Willi syndrome (PWS): Clinical and genetic comparison. Orphanet J Rare Dis 13, 1–9 (2018).

14. Barclay, S.F., et al. Absence of mutations in HCRT, HCRTR1 and HCRTR2 in patients with ROHHAD. Respir Physiol Neurobiol 221, 59–63 (2016).

15. Cielo, C. and Marcus, C.L. Central Hypoventilation Syndromes. Sleep Med Clin. 9, 105–118 (2014).

16. Barclay, S.F., et al. Rapid-Onset Obesity with Hypothalamic Dysfunction, Hypoventilation, and Autonomic Dysregulation (ROHHAD): exome sequencing of trios, monozygotic twins and tumours. Orphanet J Rare Dis 10, 103 (2015).

17. Patwari, P.P., Rand, C.M., Berry-Kravis, E.M., Ize-Ludlow, D., Weese-Mayer, D.E. Monozygotic twins discordant for ROHHAD phenotype. Pediatrics 128, e711–5 (2011).

18. Rand, C., et al. Rapid-Onset Obesity With Hypothalamic Dysfunction, Hypoventilation, and Autonomic Dysregulation: Analysis of Hypothalamic and Autonomic Candidate Genes. Pediatr Res 70, 375–378 (2011).

19. Thaker, V.V., et al. Whole exome sequencing identifies RAI1 mutation in a morbidly obese child diagnosed with ROHHAD syndrome. J Clin Endocrinol 100, 1723–30 (2005).

20. Harvengt, J., et al. ROHHAD(NET) Syndrome: Systematic Review of the Clinical Timeline and Recommendations for Diagnosis and Prognosis. J. Clin. Endocrinol. Metab. 105, 2119–2131 (2020).

21. Zaidi, S., Gandhi, J., Vatsia, S., Smith, N.L., Khan, S.A. Congenital central hypoventilation syndrome: An overview of etiopathogenesis, associated pathologies, clinical presentation, and management. Auton Neurosci. 210, 1–9 (2018).

22. Meena, J.P., et. al. Neuroblastoma presenting as opsoclonus-myoclonus: A series of six cases and review of literature. J Pediatr Neurosci. 11, 373–377 (2016).

23. Rudnick, E., et al. Opsoclonus-myoclonus-ataxia syndrome in neuroblastoma: Clinical outcome and antineuronal antibodies-a report from the children’s cancer group study. Med Pediatr Oncol. 36, 612–22 (2001).

24. Gharial, J. et al.. Neuroimaging and Pathology Findings Associated With Rapid Onset Obesity, Hypothalamic Dysfunction, Hypoventilation, and Autonomic Dysregulation (ROHHAD) Syndrome. J. Pediatr. Hematol. Oncol. 43, e571–e576 (2020).

25. Nunn, K., Ouvrier, R., Sprague, T., Arbuckle, S., Docker, M. Idiopathic Hypothalamic Dysfunction: A Paraneoplastic Syndrome? Journal of Child Neurology 12, 276–281 (1997).

26. Cemeroglu, A.P., Eng, D.S., Most, L.A., Stalsonburg, C.M., Kleis, L. Rapid-onset obesity with hypothalamic dysfunction, hypoventilation, and autonomic dysregulation syndrome and celiac disease in a 13-year-old girl: Further evidence for autoimmunity? J Pediatr Endocrinol Metab 29, 97–101 (2016).

27. Sirvent, N., et al. Hypothalamic dysfunction associated with neuroblastoma: evidence for a new Paraneoplastic syndrome? Med Pediatr Oncol 40, 326–8 (2003).

28. Sartori, S., et al. Intrathecal synthesis of oligoclonal bands in rapid-onset obesity with hypothalamic dysfunction, hypoventilation, and autonomic dysregulation syndrome: new evidence supporting immunological pathogenesis. J Child Neurol 29, 421–5 (2014).

29. Armangue, T., Petit-Pedrol, M., Dalmau J. Autoimmune encephalitis in children. J Child Neurol. 27, 1460–9 (2012).

30. Paz-Priel, I., Cooke, D.W., Chen, A.R. Cyclophosphamide for rapid-onset obesity, hypothalamic dysfunction, hypoventilation, and autonomic dysregulation syndrome. J Pediatr 158, 337–9 (2011).

31. Huppke, P., Heise, A., Rostasy, K., Huppke, B., Gärtner, J. Immunoglobulin therapy in idiopathic hypothalamic dysfunction. Pediatr Neurol 41, 232–4 (2009).

32. Voltz, R. Paraneoplastic neurological syndromes: an update on diagnosis, pathogenesis, and therapy. Lancet Neurol. 1, 294–305 (2002).

33. Dalmau, J., and Graus, F. Antibody-Mediated Encephalitis. New Engl J Med 378, 840–851 (2018).

34. Giacomozzi, C., et al. Anti-Hypothalamus and Anti-Pituitary Auto-antibodies in ROHHAD Syndrome: Additional Evidence Supporting an Autoimmune Etiopathogenesis. Horm.Res. Paediatr. 92, 124–132 (2019).

35. Rathore, G.S., Thompson-Stone, R.I., Benson, L. Chapter 20: Rapid-Onset Obesity with Hypothalamic Dysfunction, Hypoventilation, and Autonomic Dysregulation (ROHHAD). In Waubant E, Lotze TE, eds: Pediatric Demyelinating Diseases of the Central Nervous System and Their mimics: A Case-Based Clinical Guide Edition 1. Springer International Publishing 159-16 (2017).

36. Dalmau, J. and Posner, J.B. Neurologic paraneoplastic antibodies (anti-Yo; anti-Hu; anti-Ri). The case for a nomenclature based on antibody and antigen specificity. Neurology 44, 2241 (1994).

37. Vanda A. Lennon. The case for a descriptive generic nomenclature Clarification of immunostaining criteria for PCA-1, ANNA,-1, and ANNA-2 autoantibodies. Neurology 44, 2412 (1994).

38. Darnell, R.B. & Posner, J.B. Paraneoplastic Syndromes Involving the Nervous System. N Engl J Med. 16; 349, 1543–1554 (2003).

39. Ances, B.M., et. al. Treatment-responsive limbic encephalitis identified by neuropil antibodies: MRI and PET correlates. Brain 128, 1764–1777 (2005).

40. Lai, M., et. al. AMPA receptor antibodies in limbic encephalitis alter synaptic receptor location. Ann Neurol 65, 424–434 (2009).

41. O’Donovan, B., et al. High-resolution epitope mapping of anti-Hu and anti-Yo autoimmunity by programmable phage display, Brain Communications, 3;2, fcaa059 (2020).

42. Mandel-Brehm, C., et al. Kelch-like Protein 11 Antibodies in Seminoma-Associated Paraneoplastic Encephalitis. N Engl J Med. 4; 381(1), 47–54 (2019).

43. Vazquez, S.E., et al. Identification of novel, clinically correlated autoantigens in the monogenic autoimmune syndrome APS1 by proteome-wide PhIP-Seq. Elife 9, e55053 (2020).

44. Ruiz-García, R., Martínez-Hernández, E., Saiz, A., Dalmau, J., Graus, F. The Diagnostic Value of Onconeural Antibodies Depends on How They Are Tested. Front Immunol. 11, 1482. (2020).

45. Luchi. S. Three classes of C2H2 zinc finger proteins. CMLS, Cell Mol. Life Sci. 58, 625–635 (2001).

46. Dehal, P., et.al. Human chromosome 19 and related regions in mouse: conservative and lineage-specific evolution. Science 293(5527): 104–11. (2001)

47. Edelstein, L.C. & Collins, T. The SCAN domain family of zinc finger transcription factors. Gene 10, 359:1-17 (2005).

48. Sander, T.L., et al. The SCAN domain defines a large family of zinc finger transcription factors. Gene 310, 29–38 (2003).

49. Zhao, Y., et. al. RAPSearch2: a fast and memory-efficient protein similarity search tool for next-generation sequencing data. Bioinformatics. 28, 125–6 (2012).

