## Supplemental Figures, Tables and Methods for "Autoimmune profiling suggests paraneoplastic etiology in pediatric ROHHAD"

**SUPPLEMENTAL APPENDIX:**

**Supplemental Figures:**

Supplemental Figure 1: Illustrative case of ROHHAD-6 with immunotherapy

Supplemental Figure 2: Negative autoantibody detection in fixed rodent tissue

Supplemental Figure 3: Peptide-level PhIP-Seq Analysis

Supplemental Figure 4: Additional testing of ROHHAD CSF in 293T assays: Immunocytochemistry

Supplemental Figure 5: Additional testing of ROHHAD Sera in 293T assays: Immunocytochemistry

Supplemental Figure 6: Additional testing of ROHHAD CSF/Sera in 293T assays: slot-blot Western Blot

**Supplemental Tables:**

Supplemental Table 1: Autoimmune features for ROHHAD cohort

Supplemental Table 2: Expanded Neurologic and Pulmonary features for ROHHAD cohort

Supplemental Table 3: Clinical demographics for control cohort CC1

Supplemental Table 4: Clinical details for pediatric controls (OMS +/- NT, Obesity +NT)

Supplemental Table 5: Mass spectrometry analysis

**Supplemental Text:**

Supplemental Methods

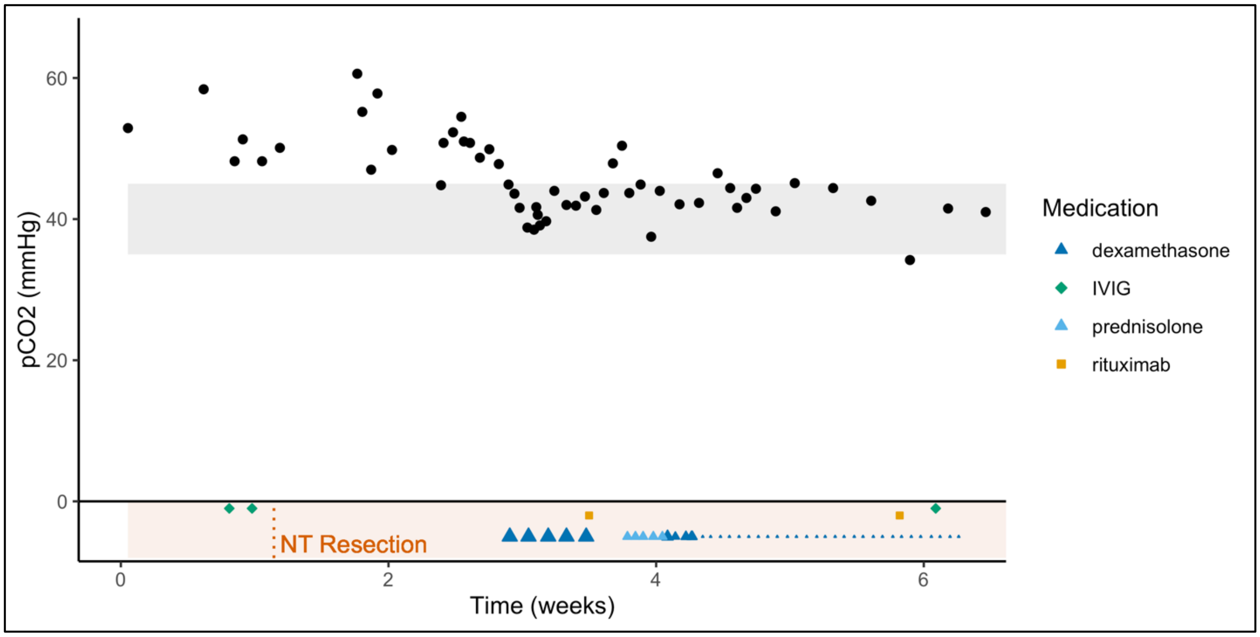


**Supplemental Figure 1. Illustrative case of improvement in ROHHAD-6 symptoms with immunotherapy.** ROHHAD-6 was admitted for asymptomatic hypernatremia in the setting of 2 years of rapid weight gain. Found to have central hypothyroidism, diabetes insipidus and day and night hypoventilation and hypoxemia requiring BiPAP initially day and night with oxygen. He was treated with IVIG 2g/kg over 2 days followed by tumor resection. Following a period of surgical wound healing he was treated with 5 days of IV dexamethasone 20mg/m2 correlated with a dramatic reduction in CO2 retention without other concurrent intervention. Rituximab 750mg/m2 first dose given on last day of pulse dexamethasone and 14 days later. CO2 rose within 2 days of stopping 5 day steroid course and improved again with resumption of IV dexamethasone after 3 days off IV steroid. Dexamethasone subsequently tapered IV then converted to PO for completion of a 16 week taper. Polysomnogram improved from “severe 24 hours hypoventilation” with peak $CO_2$ value of 60mmHg prior to any intervention to CO2 values < 50 mmHg at end of initial steroid pulse. He is maintained on rituximab to maintain B cells of 0 and IVIG 1g/kg every 4 weeks without development of additional ROHHAD symptoms or adverse event for 18 months.  With treatment he showed significant improvement in hypoventilation from severe hypoventilation requiring 24 hour respiratory support to no daytime hypoventilation with very mild snoring and mild hypoventilation on follow up $CO_2$ between 48-54mmHg requiring only BiPAP during nocturnal sleep.  We believe a good response to immunotherapy prevented the need for tracheostomy in this case.

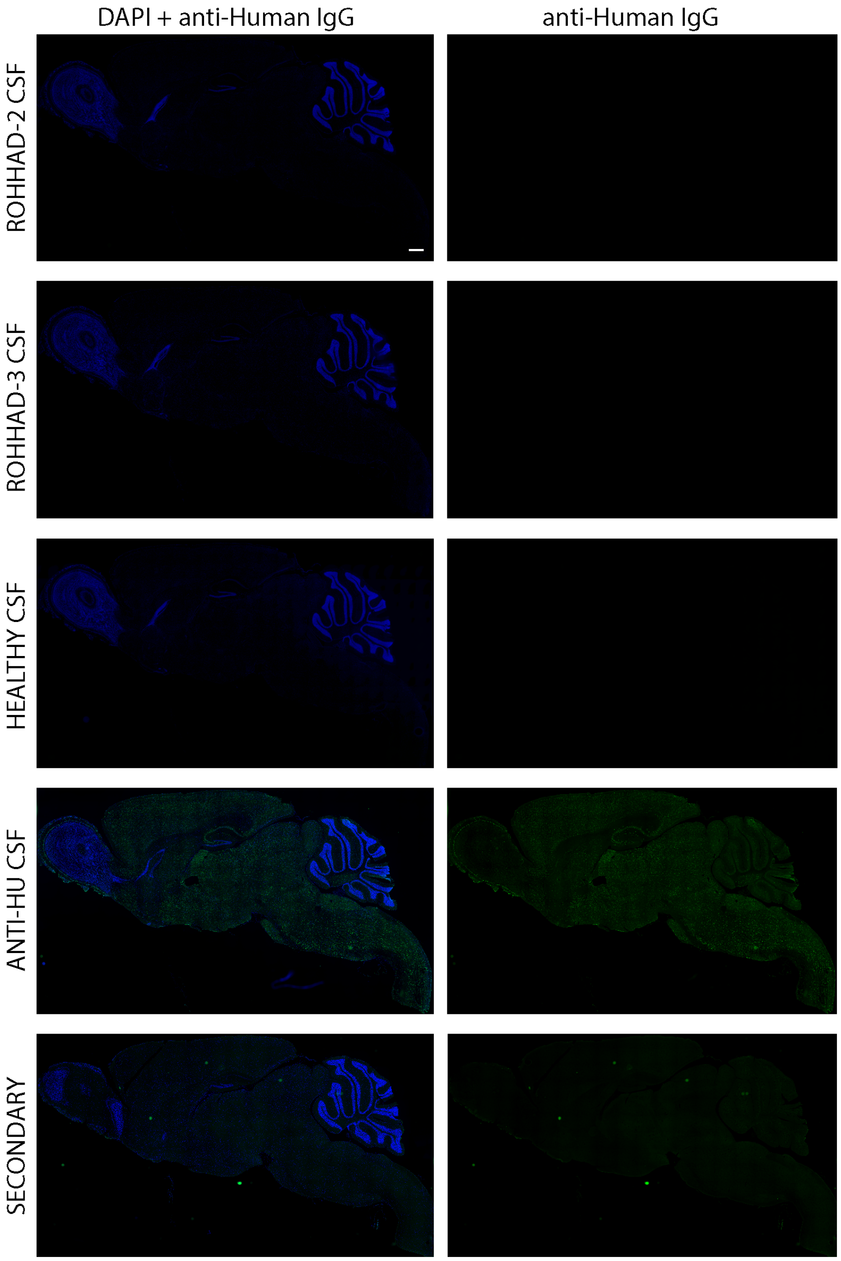


**Supplemental Figure 2. Indirect immunofluorescence on fixed sagittal mouse brain tissue.** CSF samples from ROHHAD patients 1 through 6 were screened via immunofluorescence on mouse brain tissue. Representative images of sagittal sections of select patients and controls are displayed. CSF sample from a patient identified with anti-Hu associated encephalitis[41] was used as a positive control. Blue = DAPI, Green = anti-Human IgG, scale bar represents 500 μm.

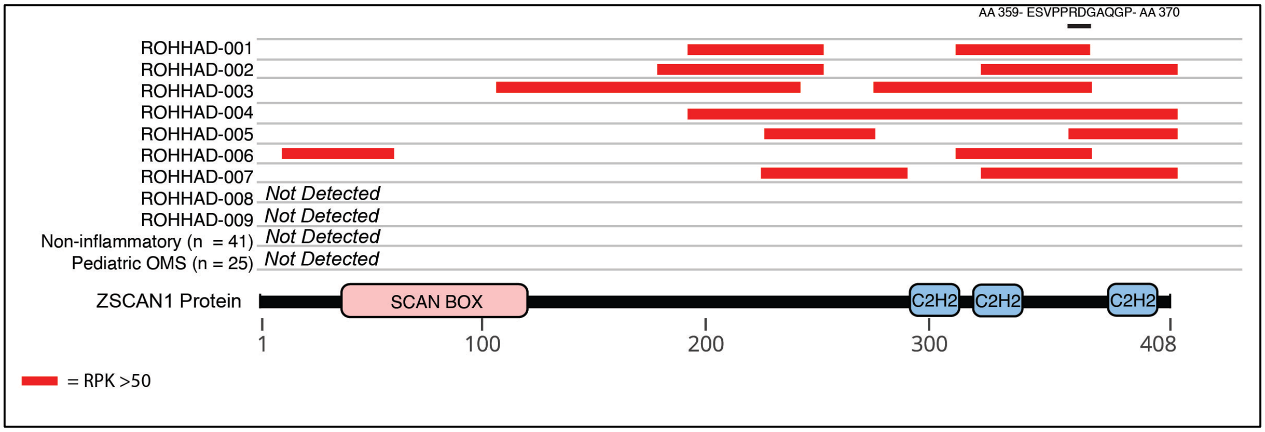


**Supplemental Figure 3. Peptide-level ZSCAN1 enrichments by ROHHAD patients, informed by PhIP-seq**. Cartoon graphic of the 408 amino acid ZSCAN1 protein with annotated SCAN and $C_2H_2$ domains is depicted below. Horizontal tracks above ZSCAN1 represent peptide enrichment data from individual ROHHAD patients or aggregated data from control cohorts. All peptides belonging to ZSCAN1 with enrichment RP100K >50 were plotted as red bars, merging together peptides with overlapping regions within individual tracks to reflect span of antigenic area. The black bar above all tracks represents an 11 AA region of overlap in 100% (7/7) patients within the C-terminal domain. The amino acid sequence for the region of overlap is depicted above the black bar.

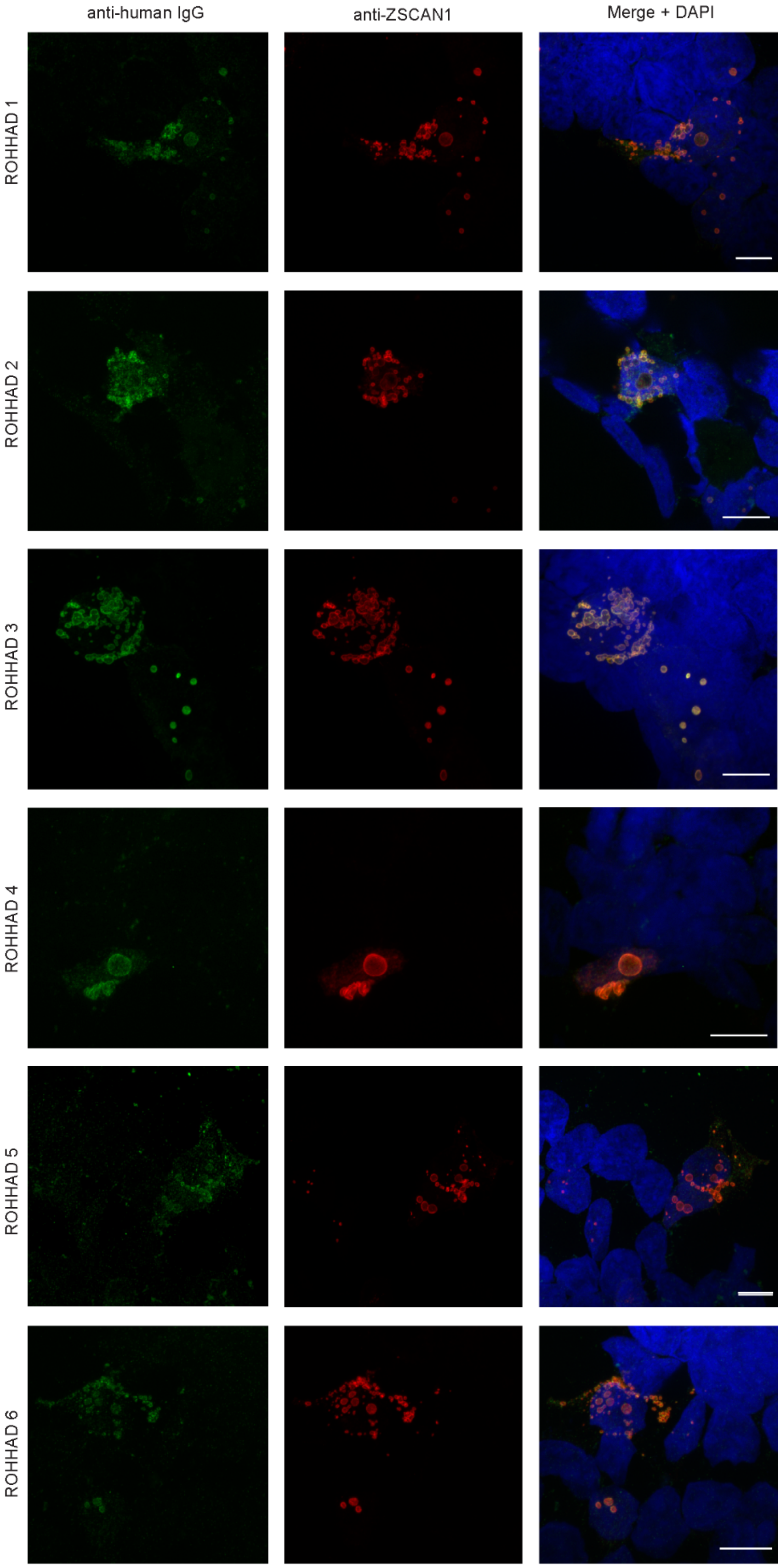
anti-human IgG
anti-ZSCAN1
Merge + DAPI
ROHHAD 1
ROHHAD 2
ROHHAD 3
ROHHAD 4
ROHHAD 5
ROHHAD 6

**Supplemental Figure 4. Validation of ZSCAN1 autoantibodies in CSF of ROHHAD patients using 293T Cell-Based Assays.** Immunocytochemistry with 293T cells expressing full-length ZSCAN1 and immunostaining with CSF (1:10) from ROHHAD patients and commercial antibody to ZSCAN1 (Rabbit, 1:1000 Invitrogen). Anti-human IgG-488 was used to visualize human IgG and anti-Rabbit IgG-567 was used to visualize anti-ZSCAN1 commercial antibody. Exposure times and post-image processing and thresholding was kept constant across conditions within the experiment. Co-localization was assessed qualitatively through observance of yellow in merged images.

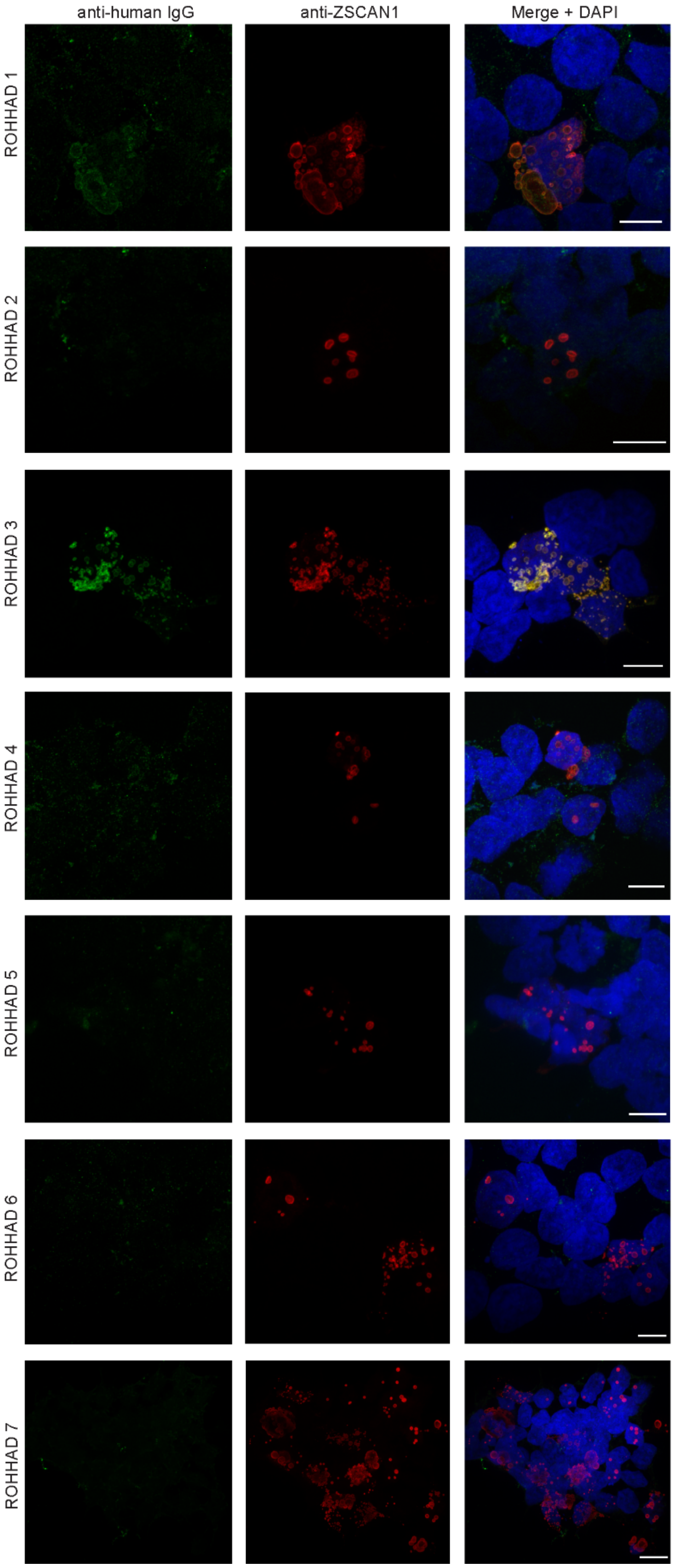
anti-human IgG
anti-ZSCAN1
Merge + DAPI
ROHHAD 1
ROHHAD 2
ROHHAD 3
ROHHAD 4
ROHHAD 5
ROHHAD 6
ROHHAD 7

**Supplemental Figure 5. Validation of ZSCAN1 autoantibodies in sera of ROHHAD patients using 293T Cell-Based Assays**. Immunocytochemistry with 293T cells expressing full-length ZSCAN1 and immunostaining with Sera (1:100) from ROHHAD patients and commercial antibody to ZSCAN1 (Rabbit, 1:1000 Invitrogen). Anti-human IgG-488 was used to visualize human IgG and anti-Rabbit IgG-567 was used to visualize anti-ZSCAN1 commercial antibody. Exposure times and post-image processing and thresholding was kept constant across conditions within the experiment. Co-localization was assessed qualitatively through observance of yellow in merged images. Note colocalization in ROHHAD Sera-1 and 3.

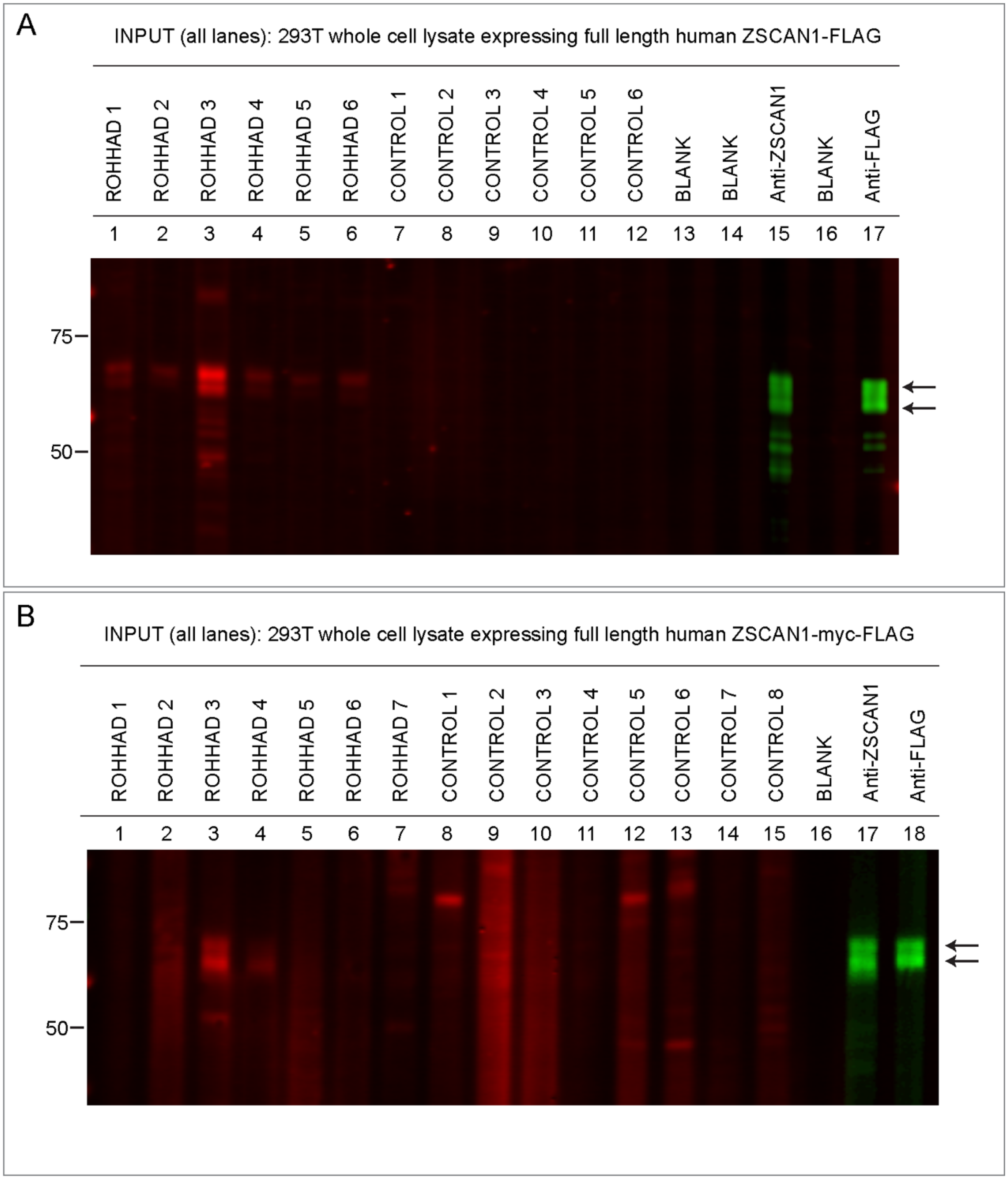


**Supplemental Figure 6. Detection of ZSCAN1 antibodies with slot-blot western blotting using ROHHAD CSF and Sera***.* Whole cell-lysates from HEK293T cells expressing transfected with full-length human ZSCAN1 cDNA were separated on a 1-well 4-12% Tris-HCl protein gel, transferred to a PVDF membrane and immunoblotted in a slot-blot device (BioRad). Primary and secondary antibodies were added to lanes as indicated. **a,** Testing ROHHAD CSF. Primary antibodies were loaded in lanes 1-12. Lanes 13 and 15 served as secondary-only controls. Primary antibodies are as follows: Lanes 1 through 7: ROHHAD 1-7 CSF (1:200); Lane 8 through 12: Controls 1-5 CSF (1:200), Lane 13: Blank, Lane 14: Commercial antibody to ZSCAN1 (Sigma, Rabbit, 1:2000); Lane 15: Blank, Lane 16: Commercial antibody to Flag (CST, Rabbit, 1:2000). Secondary antibodies to visualize human IgG Lanes 1-12: Goat anti-human IgG (LICOR680). Secondary antibodies to visualize commercial antibodies to ZSCAN1 and FLAG Lanes 14 and 16: goat anti-Rabbit IgG (LICOR800). **b,** Testing ROHHAD Sera. Primary antibodies are as follows: Lanes 1-7: ROHHAD 1-7 sera, Lane 8-15: Control 01-08, Lane 17: Commercial antibody to ZSCAN1 (Sigma, Rabbit, 1:2000), Lane 18: Commercial antibody to FLAG (CST, Rabbit, 1:2000). Secondary antibodies to visualize human IgG lanes 1-15: Goat anti-human IgG (LICOR680). Secondary antibodies to visualize commercial antibodies to ZSCAN1 and FLAG lanes 17-18: goat anti-rabbit IgG (LICOR800).

| I.D. | Diagnosis | ZSCAN1 | Autoimmunity co-morbidity | Tumor inflammation (based on clinical pathologist report) | Onset of ROHHAD symptoms following infection | Immune Modulatory and Initial Supportive Treatments (indication) | Response to Treatment | Nature of Treatment Response | Research Antibody Testing (Dalmau) |
|---|---|---|---|---|---|---|---|---|---|
| 1 | ROHHAD-NT | + | No | Extensive lymphocytic infiltrate throughout with lymphoid aggregates demonstrating germinal center formation | No | Tumor resection, IVIG, sIVIG, trach/vent | Partial, definite improvement | Enuresis resolved, fatigue, hyperphagia, temperature & heart rate stability improved | Negative |
| | | | | | | Rituximab | Possible improvement | Period of weight loss, energy improved, off vent daytime | |
| | | | | | | MMF, sIVIG | No clear improvement | | |
| | | | | | | Rituximab, sIVIG (weight gain, fatigue, delayed puberty, adrenal insufficiency and sodium variability) | Stable | No new ROHHAD symptoms | |
| | | | | | | sIVIG | Stable | | |
| 2 | ROHHAD-NT | + | No | Inflammatory cells reported within the tumor | No | Tumor resection, IVIG, sIVIG, CPAP | Improved | Weight loss, resolution of apnea | Negative |
| | | | | | | sIVIG increased to every 3 weeks for recurrence of nocturnal hypoventilation, weight gain, prolactin elevation) | No clear change/stable | | |
| | | | | | | Rituximab (ongoing weight gain, new diagnosed endocrinopathies, new daytime hypercarbia with walk test) | Possible improvement | Possible/transient improved CO2 sensitivity, weight stability | |

| | | | | | | | | | |
|---|---|---|---|---|---|---|---|---|---|
| 3 | ROHHAD-NT | + | Likely anti-phosphlipid antibody syndrome with venous sinus thrombosis and strokes (died prior to confirmatory labs) | Chronic inflammatory infiltrate (large numbers of plasma cells, lymphocytes) within the tumor. | No | Tumor resection, IVIG, sIVIG, trach/vent | Improved | Progressed from completely apneic to day time sprints off vent, to off all day after discharge; diaphoresis, constipation initially resolved; appetite reduced) | Negative |
| | | | | | | Rituximab | Unclear, response confounded by multifactorial issues and interventions. | OCBs reduced over time | |
| 4 | ROHHAD-NT | + | No (hypogammaglobulinemia) | Aggregates of lymphocytes are prominent throughout the tumor | No | Tumor resection, trach/vent, methylprednisolone, rituximab, IVIG, sIVIG, | Improved | Peak improvement 6 months after w/ weight loss, improved behavior, bowel motility, autonomic storms, resolution of diaphoresis & bathomophobia. During follow up weaned from 24 hour ventilation to day no CSA or hypoventilation, night CSA and hypoventilation requiring only nocturnal ventilator. | NA |
| | | | | | | Rituximab 2nd cycle | No clear improvement | | |
| 5 | ROHHAD-NT | + | ITP, celiac disease | NR | Onset of hyperphagia within days of rhinovirus and enterovirus URI | Tumor resection, trach/vent, dexamethasone, IVIG, sIVIG, rituximab | Minimal improvement | | NA |
| | | | | | | HiCy | Potential transient improved | Period of improved behavior, affect, stability | |
| | | | | | | Rituximab | No clear improvement | | |
| 6 | ROHHAD-NT | + | No | NR | No | Tumor resection, BiPAP, dexamethasone, IVIG, sIVIG, rituximab | Dramatic response to steroids | Direct improvement in hypercarbia | NA |

| | | | | | | | | | |
|---|---|---|---|---|---|---|---|---|---|
| | | | | | | Rituximab, sIVIG | Improved, stable | Essentially resolved central hypoventilation, resolved dysautonomia, no new manifestations | |
| 7 | ROHHAD-NT | + | No | NR | NR | Rituximab | Transient response | | NA |
| | | | | | | HiCy | Improved | Improved hyperphagia, social skills, pain perception, nocturnal saturation and hypoventilation) (Off treatment years later developed OSA) | |
| 8 | ROHHAD-NT | - | Antibody + myasthenia gravis , autoimmune thrombocytopenia (ITP) (autoimmune neutropenia, suspected EMRN | NR | No | Classic ROHHAD phenotype without immunotherapy | | | Negative |
| | | | | | | Steroids, IVIG, rituximab | Incomplete response | | |
| | | | | | | Thymectomy | Improved | Improved medical stability | |
| 9 | ROHHAD | - | No | NA | Febrile gastroenteritis preceding onset of weight gain | IVIG x1 | No clear improvement | | Negative |

**Supplemental Table 1 –Autoimmune features in the ROHHAD cohort**

AchR = Acetylcholine Receptor, BiPAP= Bilevel Positive Airway Pressure, CSF = Cerebrospinal Fluid, CPAP= Continuous Positive Airway Pressure, CSA= central sleep apnea, EMRN = Encephalomyelo-radiculo-neuropathy, IVIG = Intravenous Immunoglobulin, OCB = Oligoclonal Band, MMF = Mycophenolate Mofetil, OSA = Obstructive Sleep Apnea, NR = Not recorded, MG = Myasthenia Gravis, trach=tracheostomy, vent = ventilator

| | | | Neurologic features | | | | |
|---|---|---|---|---|---|---|---|
| **Patient No.** | **Diagnosis** | **ZSCAN1** | **Seizures** | **Behavior Problems** | **Developmental Abnormalities** | **Rumination Syndrome** | **Other complications** |
| 1 | ROHHAD-NT | + | No | No | Cognitive regression, mild ID | No | |
| 2 | ROHHAD-NT | + | No | No | No, normal neuropsych testing | No | |
| 3 | ROHHAD-NT | + | Yes (initially hyponatremia provoked) | Severe aggression, impulsivity | None early childhood, progressively delayed after disease onset, autistic features | Severe, required fundoplication | hypocalcemic tetany, extrapontine myelinolysis |
| 4 | ROHHAD-NT | + | Yes (arrest provoked) | Hyperactivity, impulsivity, agression, bathmophobia, anxiety | None early childhood, progressively delayed after disease onset/arrest | No | persistent cortical hyperintensities noted on MRI, suggestive of parenchymal injury (without hypothalamic lesions) post arrest |
| 5 | ROHHAD-NT | + | Yes | Severe aggression, impulsivity, possible psychosis | none early childhood, progressively delayed after disease onset, pain insensitivity | Yes, erythromycin helped | |

| | | | | | | | |
|---|---|---|---|---|---|---|---|
| 6 | ROHHAD-NT | + | No | No | No | No | |
| 7 | ROHHAD-NT | + | Yes | Flat affect, social withdrawl, pain insensitivity | Intellectual math disability, weaknesses in fine motor dexterity, spatial skills, verbal reasoning skills, processing speed, , pain insensitivity | No | strabismus, intermittent papular rash |
| 8 | ROHHAD-NT | - | Yes | Mood disorder | Progressively delayed after disease onset/arrest | No | Paroxysmal atrial fibrilation (pacemaker) |

| | | | | | | | |
|---|---|---|---|---|---|---|---|
| 9 | ROHHAD | - | Yes (hypernatremia provoked) | Flat affect, lethargy, pain insensitivity | No | No | MRI w/ multiple regions of scattered white matter hyperintensity, medial frontal and medial temporal gyral swelling with T2 hyperintensity bilaterally in the setting of seizures likely provoked by sodium shifts |

**Supplemental Table 2 – Additional demographics, neurologic features and genetic testing in the ROHHAD cohort**
CMA = chromosomal microarray, CSA= central sleep apnea, EMRN-encephalomyeloradiculoneuropathy (combined central and peripheral demyelination), HiCy = high dose cyclophosphamide, IBD= inflammatory bowel disease, ID= intellectual disability, IVIG = intravenous immunoglobulin, sIVIG = monthly IVIG every 3 or 4 weeks, MGM = maternal grandmother, NR= not recorded or data missing, NA = not applicable or not performed, PGM = paternal grandmother, PWS = Prader Willi syndrome, trach=tracheostomy, URI= upper respiratory infection, VUS = variant of unknown significance, WES= whole exome sequencing

| SAMPLE ID | SEX | RACE | SAMPLE ID | SEX | RACE |
|---|---|---|---|---|---|
| Healthy Plasma -1 | M | White | Healthy Plasma -51 | M | White |
| Healthy Plasma -2 | M | White | Healthy Plasma -52 | F | White |
| Healthy Plasma -3 | F | White | Healthy Plasma -53 | F | White |
| Healthy Plasma -4 | F | Hispanic/Latino | Healthy Plasma -54 | M | White |
| Healthy Plasma -5 | M | Hispanic/Latino | Healthy Plasma -55 | M | White |
| Healthy Plasma -6 | F | White | Healthy Plasma -56 | M | White |
| Healthy Plasma -7 | M | Hispanic/Latino | Healthy Plasma -57 | M | Hispanic/Latino |
| Healthy Plasma -8 | M | White | Healthy Plasma -58 | F | White |
| Healthy Plasma -9 | F | Hispanic/Latino | Healthy Plasma -59 | M | White |
| Healthy Plasma -10 | M | White | Healthy Plasma -60 | M | White |
| Healthy Plasma -11 | F | Hispanic/Latino | Healthy Plasma -61 | F | White |
| Healthy Plasma -12 | F | Multiple Race | Healthy Plasma -62 | F | White |
| Healthy Plasma -13 | F | White | Healthy Plasma -63 | F | White |
| Healthy Plasma -14 | M | Hispanic/Latino | Healthy Plasma -64 | F | White |
| Healthy Plasma -15 | M | Hispanic/Latino | Healthy Plasma -65 | F | White |
| Healthy Plasma -16 | M | White | Healthy Plasma -66 | M | White |
| Healthy Plasma -17 | F | White | Healthy Plasma -67 | F | Multiple Race |
| Healthy Plasma -18 | F | White | Healthy Plasma -68 | M | White |
| Healthy Plasma -19 | M | Black | Healthy Plasma -69 | M | White |
| Healthy Plasma -20 | F | Hispanic/Latino | Healthy Plasma -70 | M | White |
| Healthy Plasma -21 | F | Multiple Race | Healthy Plasma -71 | F | White |
| Healthy Plasma -22 | M | White | Healthy Plasma -72 | F | Hispanic/Latino |
| Healthy Plasma -23 | M | White | Healthy Plasma -73 | M | White |
| Healthy Plasma -24 | M | Asian/Pacif.Island | Healthy Plasma -74 | M | White |
| Healthy Plasma -25 | F | White | Healthy Plasma -75 | M | White |
| Healthy Plasma -26 | F | White | Healthy Plasma -76 | M | White |
| Healthy Plasma -27 | M | White | Healthy Plasma -77 | M | White |
| Healthy Plasma -28 | F | White | Healthy Plasma -78 | M | White |
| Healthy Plasma -29 | M | Hispanic/Latino | Healthy Plasma -79 | F | White |
| Healthy Plasma -30 | F | Multiple Race | Healthy Plasma -80 | F | Ethnic Group-Other |
| Healthy Plasma -31 | M | White | Healthy Plasma -81 | F | White |
| Healthy Plasma -32 | F | White | Healthy Plasma -82 | F | White |
| Healthy Plasma -33 | F | Hispanic/Latino | Healthy Plasma -83 | M | White |
| Healthy Plasma -34 | M | White | Healthy Plasma -84 | M | White |
| Healthy Plasma -35 | F | White | Healthy Plasma -85 | M | White |
| Healthy Plasma -36 | M | White | Healthy Plasma -86 | M | White |
| Healthy Plasma -37 | F | White | Healthy Plasma -87 | F | White |
| Healthy Plasma -38 | M | Hispanic/Latino | Healthy Plasma -88 | F | Asian/Pacif.Island |
| Healthy Plasma -39 | M | White | Healthy Plasma -89 | F | Hispanic/Latino |
| Healthy Plasma -40 | M | White | Healthy Plasma -90 | M | White |
| Healthy Plasma -41 | M | White | Healthy Plasma -91 | F | White |
| Healthy Plasma -42 | M | Asian/Pacif.Island | Healthy Plasma -92 | F | White |
| Healthy Plasma -43 | F | White | Healthy Plasma -93 | F | White |
| Healthy Plasma -44 | F | White | Healthy Plasma -94 | F | White |
| Healthy Plasma -45 | M | Hispanic/Latino | Healthy Plasma -95 | M | White |
| Healthy Plasma -46 | M | White | Healthy Plasma -96 | M | White |
| Healthy Plasma -47 | F | White | Healthy Plasma -97 | F | White |
| Healthy Plasma -48 | F | White | Healthy Plasma -98 | M | White |
| Healthy Plasma -49 | F | White | Healthy Plasma -99 | M | White |
| Healthy Plasma -50 | F | White | Healthy Plasma -100 | M | White |

**Supplemental Table 3: Clinical demographics of control cohort CC1**

| Patient ID | Sex | Diagnosis | Neuroblastoma (NT)? |
|---|---|---|---|
| OMS-001 | F | Paraneoplastic opsoclonus-myoclonus ataxia syndrome (OMS) | Yes |
| OMS-002 | F | opsoclonus-myoclonus ataxia syndrome (OMS) | Yes |
| OMS-003 | F | chronic relapsing opsoclonus-myoclonus ataxia syndrome (OMS) | Yes |
| OMS-004 | F | OMS | Yes |
| OMS-005 | F | OMS | Yes |
| OMS-006 | M | OMS, behavioral and emotional disorder, learning disorder | No |
| OMS-007 | F | OMS | No |
| OMS-008 | M | OMS | No |
| OMS-009 | M | OMS | No |
| OMS-010 | F | OMS, depression, tremor | No |
| OMS-011 | M | OMS | No |
| OMS-012 | M | OMS, aggressive behavior, hypogammaglobulinemia | No |
| OMS-013 | F | OMS | No |
| OMS-014 | F | Opsoclonus myoclonus ataxia syndrome (OMS) | No |
| OMS-015 | M | Neuroblastoma, OMS | Yes |
| OMS-016 | F | ADHD, Adrenal Neuroblastoma, Intellectual Disability, OMS | Yes |
| OMS-017 | M | OMS, asthma, developmental delay, distance exotropia, refractive amblyopia, stabismic amblyopia | No |
| OMS-018 | M | OMS | No |
| OMS-019 | M | OMS | No |
| OMS-020 | F | OMS | No |
| OMS-021 | M | OMS in setting of neuroblastoma | Yes |
| OMS-022 | M | Neuroblastoma associated OMS, retroperitoneal mass, lethargy, irritability, ataxia | Yes |
| OMS-023 | F | OMS, development gait and appendicular ataxia, behavioral and sleep issues | No |
| OMS-024 | F | OMS post resection of neuroblastoma and repeat resection of ganglioneuroblastoma | Yes |

| | | | |
|---|---|---|---|
| OMS-025 | F | OMS initially diagnosed as acute cerebellar ataxia | No |
| OBESITY-NT-01 | M | Obesity, NET | Yes |

**Supplemental Table 4:** Clinical details for pediatric controls (OMS +/- NT, Obesity +NT). N/A = Not Applicable, OMS = Opsoclonus Myoclonus

| Accession | Description | ZSCAN1 (Rabbit, Thermo)-replicate 1 | Rabbit Isotype Control (Thermo) | ZSCAN1 (Rabbit, Thermo)-replicate 2 | ZSCAN1 (Rabbit, Sigma) |
|---|---|---|---|---|---|
| Q8NBB4 | Zinc finger and SCAN domain-containing protein 1 OS=Homo sapiens OX=9606 GN=ZSCAN1 PE=1 SV=2 | 294211.1563 | 0 | 1356437.336 | 255851.3262 |
| A0A2R8Y6Y7 | Succinate--CoA ligase [ADP-forming] subunit beta, mitochondrial OS=Homo sapiens OX=9606 GN=SUCLA2 PE=1 SV=1 | 222871.7061 | 0 | 466508.0425 | 305563.8555 |
| A0A024RAA7 | Adiponectin B OS=Homo sapiens OX=9606 GN=ADIB PE=4 SV=1 | 2249331.242 | 0 | 600995.7988 | 283373.9453 |
| Q9P2B4 | CTTNBP2 N-terminal-like protein OS=Homo sapiens OX=9606 GN=CTTNBP2NL PE=1 SV=2 | 347039.5234 | 0 | 493797.2383 | 1377048.866 |
| Q9C037 | E3 ubiquitin-protein ligase TRIM4 OS=Homo sapiens OX=9606 GN=TRIM4 PE=1 SV=2 | 310089.5156 | 0 | 142390.459 | 546160.5742 |
| Q96JE9 | Microtubule-associated protein 6 OS=Homo sapiens OX=9606 GN=MAP6 PE=1 SV=2 | 41690931.06 | 0 | 20711782.1 | 34878341.01 |
| O43242 | 26S proteasome non-ATPase regulatory subunit 3 OS=Homo sapiens OX=9606 GN=PSMD3 PE=1 SV=2 | 386283.5059 | 0 | 164538.8706 | 680289.9609 |
| Q9Y2A7-2 | Isoform 2 of Nck-associated protein 1 OS=Homo sapiens OX=9606 GN=NCKAP1 | 171871.3535 | 0 | 1232725.41 | 1301138.511 |
| Q9UQ03 | Coronin-2B OS=Homo sapiens OX=9606 GN=CORO2B PE=1 SV=4 | 5106207.469 | 0 | 1412266.211 | 2143109.334 |
| O94905 | Erlin-2 OS=Homo sapiens OX=9606 GN=ERLIN2 PE=1 SV=1 | 386189.4824 | 0 | 3535553.731 | 6024701.408 |
| Q96PY5-3 | Isoform 2 of Formin-like protein 2 OS=Homo sapiens OX=9606 GN=FMNL2 | 9675924.566 | 0 | 29408339.55 | 36556217.97 |
| P20700 | Lamin-B1 OS=Homo sapiens OX=9606 GN=LMNB1 PE=1 SV=2 | 1503875.09 | 0 | 2226521.619 | 1414386.656 |
| O15027-5 | Isoform 5 of Protein transport protein Sec16A OS=Homo sapiens OX=9606 GN=SEC16A | 237751.2969 | 0 | 1980094.681 | 2669904.193 |

**Supplemental Table 5: Mass Spectrometry analysis of immunoprecipitated proteins from human hypothalamus using commercial antibodies to ZSCAN1 or rabbit isotype control**

### Materials and Methods

#### Patient selection and consents

All local IRB regulations were followed at Boston Children's Hospital (protocol #09-02-0043) and University of California, San Francisco (protocol #13-12236). All patient specific data was extracted by chart review (LB, LK, ST) and samples obtained following family consent.

#### PhIP-Seq analysis

PhIP-Seq experiments were performed as previously described (Supplemental Methods)[42]. One microliter of sera or 20 microliters of CSF were used for each round of PhIP-Seq. Patient antibodies were incubated with one milliliter of Phage Display library (10^10 pfu) overnight at 4 degrees, immunoprecipitated with magnetic Protein A and Protein G beads and eluted for a subsequent round of IP. Two rounds of iterative IPs were completed for each sample prior to phage elution and sequencing. Phage sequencing datasets were uploaded to an online database and analysis pipeline called "meebo" generated in-house. The analysis pipeline includes alignment of sequencing reads to input library, conversion to peptide reads, summing of peptides according to protein ID, normalization of datasets to 100,000 reads (RP100K) and calculation of mean RP100K for each protein. To identify candidate antigens, Z-score enrichments for each protein was calculated based on mean RP100K generated from a large set of healthy controls. Z-score formula = ($protein_X$ mean $RP100K_{ROHHAD}$ – $protein_X$ mean $RP100K_{control}$)/ (Standard Deviation of $protein_X$ in controls).

**Inventory of Antibodies**

Primary antibodies include: Rabbit isotype control (Thermo Fisher Scientific, 31235), rabbit-anti-Flag (Cell Signaling Technology, 14793S), rabbit anti-ZSCAN1(Thermo Fisher Scientific, catalog: PA552488), rabbit anti-ZSCAN1 (Sigma, catalog: HPA007938). Secondary antibodies include donkey-anti-human IgG Alexa 488 (Jackson Immunological Research, 709-546-149), goat-anti-rabbit IgG 546 (Thermo Fisher Scientific, A-11035). Commercial antibodies to ZSCAN1 and FLAG were validated through cell-based assays.

**Immunoprecipitation-Mass Spec**

*Immunoprecipitation (IP).* Frozen hypothalamic tissue was homogenized with a glass mortar and pestle on ice with cold RIPA buffer (Pierce, 25 mM Tris-HCL pH 7.6, 150 mM NaCl, 1% NP40, 1% Sodium Deoxycholate, 0.1% SDS, phosphatase inhibitor cocktail tablet), then passed through fine needle syringes. Lysates were incubated for 30 minutes at 4°C with gentle agitation, spun for 30 minutes at 16,000 x g at 4°C. Supernatants were transferred to a new tube and protein concentrations were measured (BCA kit, Pierce). For IP, lysate was diluted to 0.5 mg/mL in RIPA and one of the following was added; 1 ug commercial antibody to ZSCAN1 (Thermo Fisher Scientific, PA552488), 1 ug commercial antibody ZSCAN1 (Sigma Aldrich Cat: HPA007938), or 1ug of rabbit isotype control (Thermo Fisher Scientific 31235). Antibodies were incubated with lysate overnight at 4 degrees and IP'd with magnetic Protein A and Protein G beads (Thermo Fisher 1008D, 1009D). Beads were washed 3x with RIPA and 3x in RIPA without detergent. Protein on beads were submitted for Mass

Spectrometry analysis (see below). Positive identification of ZSCAN1 required detection in both commercial antibodies and absence from isotype. Two technical replicates were performed.

*Mass spectrometry analysis.* The protein bound magnetic beads were resuspend in 100 µL of 0.1% RapiGest SF Surfactant (Waters, Milford, MA) in 100mM ammonia bicarbonate. The proteins were reduced (5 mM dithiothreitol, 37 °C, 60 min) and alkylated (14 mM iodoacetamide, room temperature in dark, 45 min). 5 µg of trypsin was added to each sample for digestion overnight at 37 °C. Peptide-containing supernatant was separated from the magnetic beads and trifluoroacetic acid was added to the supernatant to adjust the pH to below 2. The sample was incubated at 37 °C for 30 minutes followed by centrifugation at 16,000 rcf for 10 minutes. One fourth of the supernatant was then subject to peptide desalting with underivatized polystyrene-divinylbenzene reverse phase S (RP-S) cartridges on an AssayMAP Bravo platform (Agilent, Santa Clara, California).

Desalted peptides were analyzed on a Fusion Lumos mass spectrometer (Thermo Fisher Scientific, San Jose, CA) equipped with a Thermo EASY-nLC 1200 LC system (Thermo Fisher Scientific, San Jose, CA). Peptides were separated by capillary reverse phase chromatography on a 25 cm column (75 µm inner diameter, packed with 1.6 µm C18 resin, AUR2-25075C18A, Ionopticks, Victoria Australia). Electrospray Ionization voltage was set to 1550 volts. Peptides resulting from on-bead digestion were resuspended in 10 µL of 0.1% formic acid. 2 µL was introduced into the Fusion Lumos mass spectrometer using a two-step linear gradient with 3–27 % buffer B (0.1% (v/v) formic acid in acetonitrile) for 105 min followed by 27-40 % buffer B for 15 min at a flow rate of 300 nL/min. Column temperature was maintained at 40°C throughout the procedure. Data was acquired in top speed data dependent mode with a duty cycle time of 1 s. Full MS scans were acquired in the Orbitrap mass analyzer with a resolution of 240 000 (FWHM) and m/z scan range of 375-1500 m/z. Selected precursor ions were subjected to fragmentation using higher-energy collisional dissociation (HCD) with quadrupole isolation window of 0.7 m/z, and normalized collision energy of 31%. HCD fragments were analyzed in the Ion Trap mass analyzer set to Turbo scan rate. Fragmented ions were dynamically excluded from further selection for a period of 45 seconds. The AGC target was set to 1,000,000 and 10,000 for full FTMS and ITMS scans, respectively. The maximum injection time was set to Auto for both full FTMS scans and ITMS scans.

The resulting data were searched using SEQUEST HT on the Proteome Discoverer (2.2.0.388) platform, against a database combining Human proteins (downloaded January 28, 2021) and common contaminants. The precursor mass range was set to 350-5000 Da, the mass error tolerance was set to 10 ppm, and the fragment mass error tolerance to 0.6 Da. Enzyme specificity was set to trypsin, carbamidomethylation of cysteines (+57.021) was set as fixed modifications, oxidation of methionines (15.995) and acetylation of protein N-terminus (+42.011) was considered as variable modifications. Percolator was used to filter peptides and proteins to a false discovery rate of 1%.  Abundance quantification was based on precursor ion intensities.

**293T cell-based assay: transfections**

*Tissue culture.* HEK293T cells (ATCC CRL-3216) were maintained in Dulbecco's modified Eagle's medium with glutamine, 10% Fetal Bovine Serum, 50 units of penicillin and streptomycin at 5% CO2 and 37°C. For immunocytochemistry (ICC) experiments, cells were plated in 24-well tissue culture plates on coverslips pre-washed in acetic acid and coated with poly-D-lysine (Sigma-Aldrich). For IP and western blotting, cells were plated on standard 10 $cm^2$ culture dishes. *Transfections.* 24 hours after seeding, 293T cells were transfected with experimental plasmids using the lipid-based Lipofectamine 3000 (Invitrogen). For ICC experiments, 500 nanograms of a flag-tagged human ZSCAN1 expressing plasmid (Origene, RC221074) was transfected per well. For molecular experiments, one microgram of ZSCAN1 expressing plasmid was transfected per dish.

**293T cell-based assay: Immunocytochemistry and imaging**

*Staining.* 24 hours post-transfection, cells were washed once in cold 1x PBS and fixed in 2% paraformaldehyde for 15 minutes at room temperature. Cells were stained according to standard procedure with one of the following: Patient IgG (CSF 1:100, Serum 1:1000), commercial anti-ZSCAN1 IgG (1 ug/ml) or commercial anti-FLAG IgG (1 ug/ml). (Supplemental Methods). Staining protocol: Following washing with 1x PBS, cells were permeabilized and blocked for one hour at room temperature in Blocking Buffer (1x PBS, 10% Lamb Serum, 0.5% Triton). Following blocking, cells were incubated in primary antibody buffer (1x PBS 10% Lamb Serum, 0.1% Triton) overnight at 4°C containing one of the following: Patient IgG (CSF 1:100, Serum 1:1000), commercial anti-ZSCAN1 IgG (1 ug/ml) or commercial anti-FLAG IgG (1 ug/ml). Proper controls were performed. Cells were washed in 3x PBST (1X PBS with 0.02% Triton-X) and incubated with secondary antibodies (anti-Human IgG Alexa 488, anti-rabbit IgG or anti-mouse IgG Alexa 568) for 2 hours at room temperature, protected from light. Following secondary antibody incubation, cells were washed 6x in PBST, with DAPI added to the last wash. Coverslips were mounted on glass slides using prolong gold (Invitrogen). *Imaging.* Images were captured using a Nikon Ti CSU-W1 Spinning Disk/High Speed Widefield microscope at the UCSF Nikon Core Facility with a Plan Apo 20x/0.75 or Plan Apo VC 100x/1.4 oil immersion lens. Image capture settings, including exposure time, laser intensity, aperture, and magnification, were kept constant for all conditions in the experiment and analyzed in ImageJ.

**293T cell-based assay: Slot-blot Western Blotting**

Twenty-four hours post-transfection, cells were washed once in cold 1XPBS, then harvested and lysed with RIPA buffer for 30 minutes at 4°C with gentle agitation. Lysates were spun down for 30 minutes at 16,000 x g at 4°C. Supernatants were diluted to a 1 mg/ml protein concentration using the BCA (Pierce) kit and prepared for SDS/PAGE and slot -blot western blotting (see supplemental methods). 4x laemmli buffer was added to lysates, boiled for 5 minutes at 95°C, loaded onto a 4-12% gradient SDS-PAGE gel (Bio-Rad), then transferred to a 0.20 micron PVDF membrane. Membranes were blocked at room temperature for two hours in blocking buffer (LI-COR), then loaded onto a multiscreen apparatus to screen for multiple CSF or serum samples as well as commercial antibodies. Human CSF samples were loaded at 1:200, human serum samples were loaded at 1:10000, while commercial antibodies were loaded at 1:2000 and incubated overnight at 4°C. To visualize proteins of interest, anti-Human IgG (LI-COR 680) and anti-rabbit IgG (LI-COR 800) were loaded into wells and incubated for 2 hours at room temperature and scanned on Odyssey imaging system (LI-COR).

**Indirect immunofluorescence assay and microscopy on fixed mouse brain tissue.** Adult C57B6 mice were perfused with 4% PFA and the brain was dissected and cryopreserved in 30% sucrose. Brains were embedded in OCT via slow freezing on dry ice and stored at -80°C. Brain sections were collected using a cryostat at 15 microns onto superfrost plus microscope slides. Brain sections were stored at -20°C. Tissue sections were blocked and permeabilized in blocking buffer (0.2% Triton-X, 10% Lamb Serum, 1X PBS). Following blocking, sections were incubated overnight at 4°C in primary antibody buffer with ROHHAD patient CSF or a positive control (Anti-Hu) (CSF 1:10). Tissues were washed in washing buffer (1X PBS, 0.02% Triton-X) and incubated in secondary antibodies for 1 hour at room temperature, protected by light. To detect human antibodies, anti-Human IgG Alexa-488 (Jackson) was used. Tissue sections were washed and mounted using Prolong Gold Anti-Fade Reagent (Invitrogen). Experiments were visualized and image tiles were stitched using a ZEISS Axioscan 7 and a ZEISS Axio Imager 2. Image capture settings, including exposure time, laser intensity, aperture, and magnification were kept constant for all conditions.

**Radioligand Binding Assay (RLBA)**

The RLBA was performed as described previously.[43] Briefly, full-length human ZSCAN1 was in vitro transcribed and translated with [35S]-methionine in a T7 dependent system using a ZSCAN1 plasmid containing a T7 promoter (Origene Cat: RC221074). The radiolabeled ZSCAN1 protein was then column purified and immunoprecipitated with 2.5ul serum or CSF per well using Sephadex protein A/G beads (Sigma Aldrich, St. Louis, MO; #GE17-5280-02 and #GE17-0618-05) in 96-well filtration plates (Corning, Corning, NY; #EK-680860). The plates were read out as counts per minute (cpm) using the Microbeta Trilux liquid scintillation plate reader (Perkin Elmer).
